# Ethnic differences of genetic risk and smoking in lung cancer: two prospective cohort studies

**DOI:** 10.1101/2023.02.09.23285130

**Authors:** Meng Zhu, Jun Lv, Yanqian Huang, Hongxia Ma, Ni Li, Xiaoxia Wei, Mengmeng Ji, Zhimin Ma, Ci Song, Cheng Wang, Juncheng Dai, Fengwei Tan, Yu Guo, Robin Walters, Iona Y. Millwood, Rayjean J. Hung, David C. Christiani, Canqing Yu, Guangfu Jin, Zhengming Chen, Qingyi Wei, Christopher I. Amos, Zhibin Hu, Liming Li, Hongbing Shen

**Author notes:** **Corresponding authors:** Hongbing Shen, Department of Epidemiology, Center for Global Health, School of Public Health, Nanjing Medical University, 101 Longmian Avenue, Nanjing 21116, China; or Liming Li, Department of Epidemiology and Biostatistics, Peking University Health Science Center, 38 Xueyuan Road, Beijing 100191, China; or Zhibin Hu, State Key Laboratory of Reproductive Medicine, Center for Global Health, Nanjing Medical University, 101 Longmian Avenue, Nanjing 21116, China. These authors contributed equally to this work (Meng Zhu, Jun Lv, Yanqian Huang, Hongxia Ma, Ni Li). Joint last authors, contributed equally (Zhibin Hu, Liming Li, Hongbing Shen).

## Abstract

**Background:** The relative risk of smoking on lung cancer have been reported to be much higher in white population than that in East Asians. However, it’s unknown whether genetic background underlies this disparity between ethnic groups. To assess the role of ethnic differences in genetic factors associated with this phenomenon.

**Methods:** We first constructed ethnic-specific polygenic risk scores (PRSs) to quantify individual genetic risk of lung cancer in Chinese and white populations. Then, we compared genetic risk and smoking as well as their interactions on lung cancer between two cohorts, including the China Kadoorie Biobank (CKB) and the UK Biobank (UKB). We also evaluated the absolute risk reduction over a 5-year period.

**Results:** 19 SNPs and 23 SNPs were identified to construct the PRSs in Chinese and white populations, and smoking-related loci were only included in white populations. The PRSs were consistently associated with lung cancer risk respectively, but stronger associations were observed in smokers of the UKB (HR 1.26 *versus* 1.15, *P*=0.028). A significant interaction between genetic risk and smoking on lung cancer was observed in the UKB (RERI, 11.39 [95% CI, 7.01-17.94]), but not in the CKB. By comparing heavy smokers with nonsmokers, a greater absolute risk reduction was found in the UKB (10.95 *versus* 7.12 per 1000 person-years, *P*<0.001), especially for those at high genetic risk.

**Conclusions:** In China, tobacco control alone is not enough to reduce the burden of lung cancer, and comprehensive policies should be made to lower its high incidence.

## Background

Lung cancer continues to be the leading cause of cancer death worldwide. Although tobacco smoking is the main risk factor for lung cancer, the reported relative risks (RRs) and absolute risks for lung cancer morbidity associated with tobacco smoking are much lower in East Asians (RRs, 2.4-6.5) than in white population (RRs, 9.4-23.2) (1-3). Several theories have been proposed to explain this phenomenon, “the smoking paradox”, but few were based on solid data, especially with regard to ethnic differences in genetic factors (4). Tobacco control efforts implemented in high income countries since the 1960s have led to considerable reductions in lung cancer (5). However, the potential impact of tobacco control on the burden of lung cancer in Chinese population still needs further evaluation.

The development of lung cancer results from an intricate interplay between genetic and environmental factors, and the heritability of lung cancer has been estimated at 15%-18% (6, 7). In the past decade, genome-wide association studies (GWASs) of lung cancer have identified 51 risk loci, most of which were derived from populations of white and East Asian (8). Recent large-scale population studies have demonstrated that the combined effect of these genetic loci, polygenic risk score (PRS), can serve as an efficient tool to quantify individual inherent risk of lung cancer in populations of white and Chinese descents (9, 10). However, the two previously reported ethnic-specific PRSs for lung cancer are not directly comparable because of the differences in underlying genetic architectures, constructing strategy, and selecting susceptibility loci.

To comprehensively explore the genetic disparity underlying “the smoking paradox”, we generated Chinese-specific and White-specific PRSs for lung cancer by using the largest available GWAS datasets of lung cancer for populations of Chinese and white with unified standards and processes. Then, we used data from two nationwide prospective cohorts, the China Kadoorie Biobank (CKB) and the UK Biobank (UKB), to evaluate the effectiveness of the PRSs in predicting lung cancer risk and to dissect the complex relations between smoking and genetic risk in Chinese and white populations, respectively.

## Methods

### Study populations

The CKB is a population-based prospective cohort study in China. The study design for the CKB has been described previously (11). In brief, a total of 512,714 Chinese adults aged 30-79 years were recruited from ten geographically diverse regions across China between 2004 and 2008. All participants completed an interviewer-administered electronic questionnaire on smoking and other health-related information, underwent physical measurements, and provided blood samples at baseline. A total of 100,641 participants were selected for genotyping based on a clustered random selection method (12). For the present study, we included individuals with both genotypic and phenotypic data and excluded those with lung cancer diagnosed before baseline, leaving 100,615 eligible participants in the final analysis.

The UKB is also a population-based cohort study, with more than 500,000 participants aged 37-73 years who were recruited from 22 centers throughout the United Kingdom between 2006 and 2010. Details of the study have been described previously (13). Each participant provided information on smoking and other health-related information through extensive baseline questionnaires, interviews, physical measurements, and a blood sample collected for genotyping. Of the 502,527 available participants, we excluded those withdrawing from the UKB, of non-white decent, with lung cancer at baseline, with missing information on genotypes and smoking, or those failed in quality control of genotypes. Overall, 406,880 eligible participants were included in the present study.

Each participant in the CKB and the UKB completed a written informed consent form. The CKB has been approved by the Ethical Review Committee of the Oxford Tropical Research Ethics Committee, University of Oxford and the Chinese Center for Disease Control and Prevention. The UKB has been approved by the multicenter Research Ethics Committee, the National Information Governance Board for Health and Social Care in England and Wales, and the Community Health Index Advisory Group in Scotland.

### Procedures

We used the largest available GWAS datasets of lung cancer in populations of Chinese (13,327 cases and 13,328 controls) (9) and of white descent (29,266 cases and 56,450 controls) (14) to evaluate the associations and corresponding effects of all the previously reported single-nucleotide polymorphisms (SNPs; e.g. 81 SNPs in 51 loci) associated with lung cancer risk. Meanwhile, a meta-analysis dataset of lung cancer with balanced sample sizes of white descent (13,793 cases and 14,027 controls from the INTEGRAL-ILCCO OncoArray Project) and Chinese descent (13,327 cases and 13,328 controls) was used to evaluate selective candidate variants to construct a trans-ancestry PRS for sensitivity analysis (9, 10). Details of these GWAS datasets have been reported elsewhere previously (9, 10, 14). The criteria to exclude the redundant variants and the process for genotyping and imputation used in the CKB and UKB are described in the Supplement.

The PRSs were created following an additive genetic model as previously described. In short, the dosage of each risk allele for each individual was summed after multiplication with its respective weight (e.g. the Ln of the odds ratio [OR]) derived from the datasets mentioned above. The processes of PRS construction were blinded to the endpoints in the CKB and the UKB.

Smoking measures were self-reported at initial assessment with an interviewer-administered questionnaire in the CKB or a touchscreen in the UKB (11, 13). Based on pack years smoked, participants were defined as nonsmokers (less than 100 cigarettes in lifetime), light smokers (pack-years<30), and heavy smokers (pack-years≥30). The assessment of covariates is provided in the **online data supplement**.

The primary outcome for analysis was the event of incident lung cancer, classified by the 10th Revision of International Classification of Diseases (ICD-10 codes C33-34). Participants in the CKB cohort were followed up through ongoing electronic linkage with the Chinese national health insurance claim database, established chronic disease registries, and local death registries semi-annually, supplemented by active confirmation of cancer diagnosis by trained staff. Complete follow-up for the CKB was available through December 31, 2016. For the UKB, incident lung cancer events were ascertained through linkage to national cancer registries in England, Wales and Scotland. The complete date of follow-up was March 31, 2016 for England and Wales, and October 31, 2015 for Scotland.

### Statistical analyses

We assessed lung cancer risk in participants from the enrolment until the time of lung cancer diagnosis, death, or the end of follow-up, whichever occurred first. We assessed a potential nonlinear relationship between the PRS and lung cancer risk by use of restricted cubic spline analysis. Cox proportional hazard models were used to assess associations between genetic factors and smoking with lung cancer incidence and to estimate hazards ratios (HRs) and 95% confidence intervals (CIs). Schoenfeld residuals were used to test the proportional hazards assumption. The genetic risk was categorized into low (the bottom quintile), intermediate (quintiles 2-4) and high (the top quintile) based on distributions of PRSs, as described previously (12). Relative excess risk due to interaction (RERI) and the attributable proportion because of the interaction (AP) were calculated to measure the interaction on the additive scale (15). The mediation proportion by the mediator was calculated by comparing estimates from models with and without the hypothesized mediator (16). We then calculated cumulative risk as the incidence of lung cancer occurring in a given group during follow-up. We also calculated absolute risk reduction as the difference in lung cancer incidence between given groups over a 5-year period.

We performed several sensitivity analyses to examine the robustness of the results: (1) trans-ancestry PRSs with the same SNPs and effects were generated simultaneously to define genetic risk of the participants; (2) additional environmental exposures were included in the model, including passive smoking and ambient PM_2.5_ concentration; (3) genetic risk levels were reclassified by quartile or tertiles; (4) smoking status was reclassified as never, former, and current smokers; (5) participants who were diagnosed with lung cancer within the first year of follow-up were excluded; and (6) analysis were restricted to participants with complete covariates for comparison with the results of imputation. All *P*-values were two-sided, and *P*<0.05 was considered statistically significant. All statistical analyses were performed in R (version 3.5). Further details about statistical analyses are provided in the **online data supplement**.

## Results

The study design is shown in **Figure E1**, and the baseline characteristics of participants are provided in **Table 1**. More smokers were observed in the UKB (45.16%) than in the CKB (34.38%). In the CKB, during a median follow-up of 10.42 years (IQR 9.34-11.30), 1,392 incident lung cancer cases were diagnosed; while there were 2,025 incident lung cancer cases in the UKB during a median follow-up of 7.17 years (IQR 6.48-7.75). In both the CKB and UKB, differences in the incidence of lung cancer were observed among nonsmokers, light smokers, and heavy smokers, while similar incidence was observed between male and female under the same smoking status (**Table E1**).

**Table 1.** Baseline characteristics of participants from the China Kadoorie Biobank and the UK Biobank

| Variables | China Kadoorie Biobank |  | UK Biobank |  |
| --- | --- | --- | --- | --- |
|  | No Lung Cancer<br>(N=99,223) | Incident Lung Cancer<br>(N=1,392) | No Lung Cancer<br>(N=404,855) | Incident Lung Cancer<br>(N=2,025) |
| Age at baseline, mean (SD), years | 53.59±10.99 | 61.79±9.04 | 56.87±8.00 | 62.12±5.62 |
| Men | 42142 (42.47) | 856 (61.49) | 185805 (45.89) | 1068 (52.74) |
| Previous cancer diagnosis | 385 (0.39) | 10 (0.72) | 26753 (6.61) | 240 (11.85) |
| Family history of cancer | 16363 (16.49) | 248 (17.82) | 145342 (35.90) | 892 (44.05) |
| Emphysema and/or bronchitis | 3705 (3.73) | 127 (9.12) | 7332 (1.81) | 206 (10.17) |
| Maximum FEV1, mean (SD), liters | 2.19±0.69 | 1.93±0.68 | 2.85±0.77 | 2.45±0.74 |
| BMI |  |  |  |  |
| -18.5 | 5087 (5.13) | 133 (9.55) | 2004 (0.49) | 27 (1.33) |
| 18.5-25 | 60702 (61.18) | 876 (62.93) | 131269 (32.42) | 621 (30.66) |
| 25-30 | 28958 (29.19) | 344 (24.71) | 173860 (42.94) | 855 (42.22) |
| 30- | 4475 (4.51) | 39 (2.80) | 97722 (24.14) | 522 (25.78) |
| Smoker | 33779 (34.04) | 809 (58.12) | 181993 (44.95) | 1736 (85.73) |
| Pack-Year, mean (SD) | 27.73±21.21 | 37.58±23.78 | 23.12±15.35 | 38.99±24.01 |
| -10 | 6500 (19.24) | 73 (9.02) | 30842 (16.95) | 88 (5.07) |
| 10-20 | 7748 (22.94) | 125 (15.45) | 59966 (32.95) | 247 (14.23) |
| 20-30 | 7020 (20.78) | 134 (16.56) | 57160 (31.41) | 416 (23.96) |
| 30- | 12511 (37.04) | 477 (58.96) | 34025 (18.70) | 985 (56.74) |
| Ethnic-specific PRS, mean (SD) <sup>†</sup> | 1.78±0.36 | 1.84±0.36 | 2.60±0.33 | 2.68±0.33 |
| Low | 19902 (20.06) | 221 (15.88) | 81103 (20.03) | 273 (13.48) |
| Intermediate | 59547 (60.01) | 822 (59.05) | 242916 (60.00) | 1212 (59.85) |
| High | 19774 (19.93) | 349 (25.07) | 80836 (19.96) | 540 (26.67) |
| Trans-ancestry PRS, mean (SD) <sup>‡</sup> | 1.84±0.31 | 1.90±0.31 | 1.91±0.34 | 1.98±0.34 |
| Low | 19888 (20.04) | 235 (16.88) | 81094 (20.03) | 287 (14.47) |
| Intermediate | 59580 (60.05) | 789 (56.68) | 242908 (59.99) | 1215 (60.00) |
| High | 19755 (19.91) | 368 (26.44) | 80853 (19.97) | 523 (25.83) |

Abbreviation: FEV1, forced expiratory volume in 1 second; PRS, polygenic risk score;
<sup>†</sup> Ethnic-specific PRS was defined as low (the bottom quintile), intermediate (quintiles 2 to 4), and high (the top quintile) according to distributions of ethnic-specific PRS in CKB and UKB cohort;
<sup>‡</sup> Trans-ancestry PRS was defined as low (the bottom quintile), intermediate (quintiles 2 to 4), and high (the top quintile) according to distributions of trans-ancestry PRS in CKB and UKB cohort.

The associations of 81 reported susceptibility loci with lung cancer in GWAS datasets of Chinese population and white population are shown in **Table E2**. After systematic evaluation and filtering, 19 SNPs and 23 SNPs were retained for the calculation of Chinese-specific (PRS-19) and White-specific PRSs (PRS-23), respectively (**Figure E2**). As shown in **Figure 1**, the majority of the susceptibility loci were ethnic specific, and differences in the frequency and association effects were also observed in shared loci between two populations. Specifically, the loci of 8p21.2-*CHRNA2* (rs11780471), 15q25.1-*CHRNA5* (rs55781567), and 19q13.2-*CYP2A6* (rs56113850) that were smoking-related (14), showed higher frequencies and greater effects in white population.

**Figure 1.**
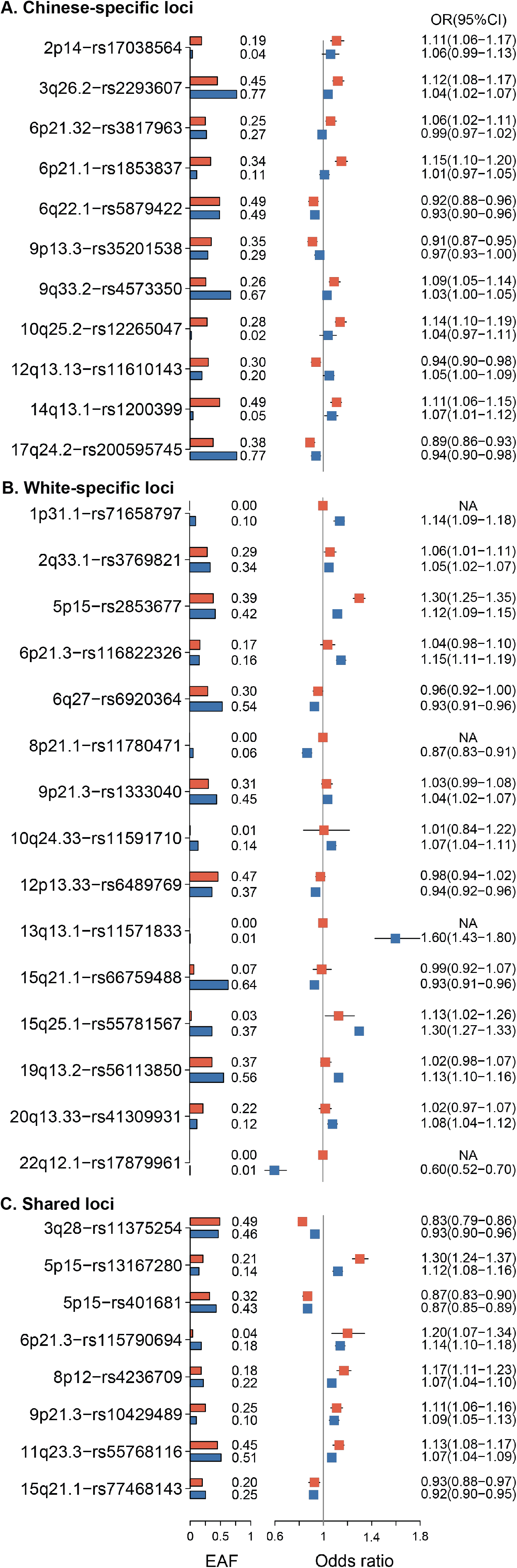
Forest plot of lead variants in the construction of Chinese-specific and European-specific polygenic risk scores (PRSs). The squares indicate ORs of analysis in Chinese or European populations. The bars are 95% CIs. MAF is indicated at the middle of the plot. The indicator of source of variants is indicated at the right of the plot.

The PRS-19 and PRS-23 were consistently associated with lung cancer risk in a linear and dose-response relationship in the CKB and UKB, respectively (**Figure E3**). Compared with participants at low genetic risk, participants at intermediate and high genetic risk had significantly greater risks of lung cancer, with HRs of 1.25 (95% CI, 1.08-1.45) and 1.62 (95% CI, 1.37-1.92) in the CKB, and 1.44 (95% CI, 1.26-1.64) and 1.87 (95% CI, 1.62-2.17) in the UKB, respectively (**Table 2**). In nonsmokers, similar linear associations and effects were observed between two cohorts (*P*_heterogeneity_=0.898); however, in smokers, greater associations were observed in the UKB than in the CKB (HR 1.26 [95% CI, 1.20-1.32] *versus* 1.15 [95% CI, 1.07-1.23] per SD of PRSs increase; *I*^2^=79.3%, *P*_heterogeneity_=0.028) (**Figure 2**). In addition, the observed associations were attenuated, if the ethnic-specific PRSs were cross-used (**Table E3**). These results did not change significantly when additional environmental exposures were included in the model, or genetic risk was reclassified by quartile or tertiles of the PRSs, or the analysis were restricted to participants without missing covariates, or by excluding incident cases occurred during the first year of follow-up (**Table E4-7**).

**Table 2.** Associations of ethnic-specific polygenic risk score (PRS) with incident lung cancer in the CKB and the UKB

| Genetic risk <sup>†</sup> | Chinese-specific PRS in the CKB |  |  | White-specific PRS in the UKB |  |  |
| --- | --- | --- | --- | --- | --- | --- |
|  | No. of cases /<br>Person-years | HR (95% CI) <sup>‡</sup> | *P*-value <sup>‡</sup> | No. of cases /<br>Person-years | HR (95% CI) <sup>‡</sup> | *P*-value <sup>‡</sup> |
| Per SD increase of PRS | 1,392/990,449 | 1.19(1.13-1.25) | $2.73 \times 10^{-10}$ | 2,025/2,875,132 | 1.24(1.19-1.29) | $7.01 \times 10^{-23}$ |
| Low | 221/198,111 | Ref |  | 273/575,454 | Ref |  |
| Intermediate | 822/594,440 | 1.25(1.08-1.45) | 0.004 | 1,212/1,724,837 | 1.44(1.26-1.64) | $6.36 \times 10^{-8}$ |
| High | 349/197,898 | 1.62(1.37-1.92) | $2.64 \times 10^{-8}$ | 540/574,841 | 1.87(1.62-2.17) | $2.94 \times 10^{-17}$ |
| *P*-value for trend | | | $1.36 \times 10^{-8}$ | | | $8.17 \times 10^{-18}$ |

<sup>†</sup> Genetic risk were categorized into low (the bottom quintile), intermediate (quintiles 2-4) and high (the top quintile) according to distributions of PRSs ;
<sup>‡</sup> Adjusting for age, sex, smoking status, BMI, highest education level, family history of cancer, personal medical history (previous cancer diagnoses and chronic obstructive pulmonary disease), the forced expiratory volume in 1 second, and the top ten principal components of ancestry.

**Figure 2.**
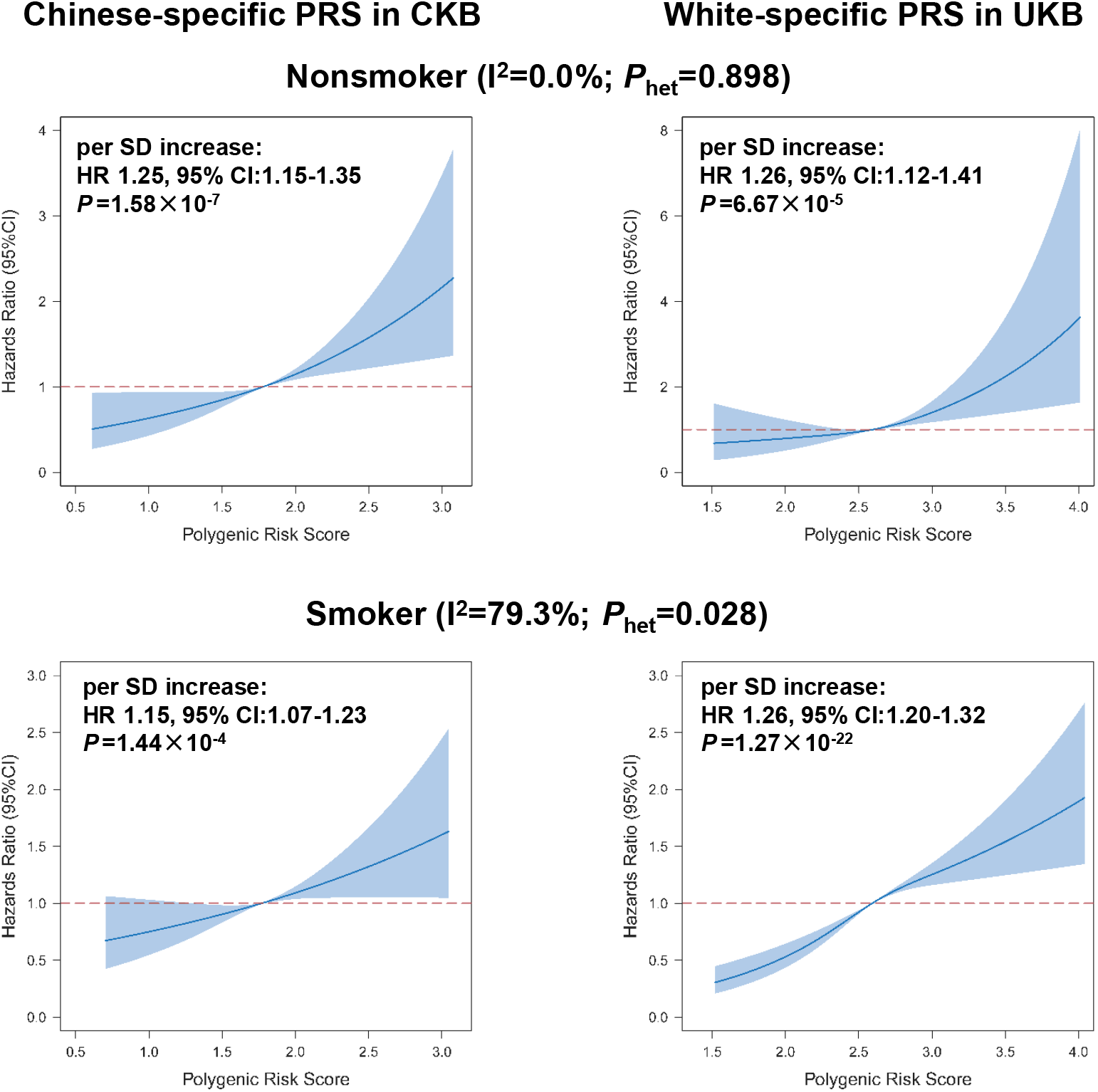
The association effects of ethnic-specific polygenic risk scores (PRSs) with incident lung cancer in CKB and UKB cohorts stratified by smoking status. The association effects between ethnic-specific PRSs and incident lung cancer risk in CKB were shown in left; and the corresponding associations in UKB were shown in right. Linear relationship between PRS and lung cancer risk was assessed using a restricted cubic spline analysis, and hazards ratios (HRs) were estimated with adjustment for age, sex, BMI, highest education level, family history of cancer, personal medical history, the forced expiratory volume in 1 second, and the top ten principal components of ancestry.

We also observed “the smoking paradox” that the relative risks were 2.87 (95% CI, 2.40-3.44) in the CKB and 15.79 (95% CI, 13.77-18.10) in the UKB among heavy smokers compared with nonsmokers. The associations did not change after further adjustment for genetic risk (**Table E8**), and similar results were observed in current smokers compared with nonsmokers (**Table E9**). The PRS-19 was not associated with smoking amount (i.e., pack-year) in the CKB, but the PRS-23 was significantly associated with smoking amount in the UKB (*P*<0.001, **Table E10**), which mediated a proportion of 2.06% (95% CI, 1.41%-2.90%) for the association between PRS-23 and incident lung cancer (**Figure E4**). To rule out the influences of different PRS compositions, we also constructed a PRS based on 25 SNPs from a trans-ancestry GWAS meta-analysis (**Figure E5**), and observed a similar mediation effect only in the UKB but not in the CKB (**Table E11**).

We further evaluated the joint effect of genetic risk and smoking on lung cancer risk and found that the HRs of participants with a high genetic risk and heavy smoking were 4.95 (95% CI, 3.61-6.77) and 25.63 (95% CI, 18.58-35.36) in the CKB and the UKB, respectively, compared with those with a low genetic risk and never smoking (**Figure 3**). Furthermore, we observed significantly additive interactions between genetic risk and smoking on incident lung cancer in the UKB but not in the CKB (**Table 3**). Specifically, for heavy smokers with a high genetic risk, the RERI was 11.39 (95% CI, 7.01-17.94), accounting for 44% (95% CI, 32%-55%) of the risk in those participants who had both a high genetic risk and heavy smoking in the UKB. We repeated the analyses by using the trans-ancestry PRS, reclassifying genetic risk levels by quartile or tertiles of the PRS, or excluding incident lung cancer occurred during the first year of follow-up, and observed similar additive interactions in the UKB but not in the CKB (**Figure E6** and **Table E12-16**).

**Table 3.**
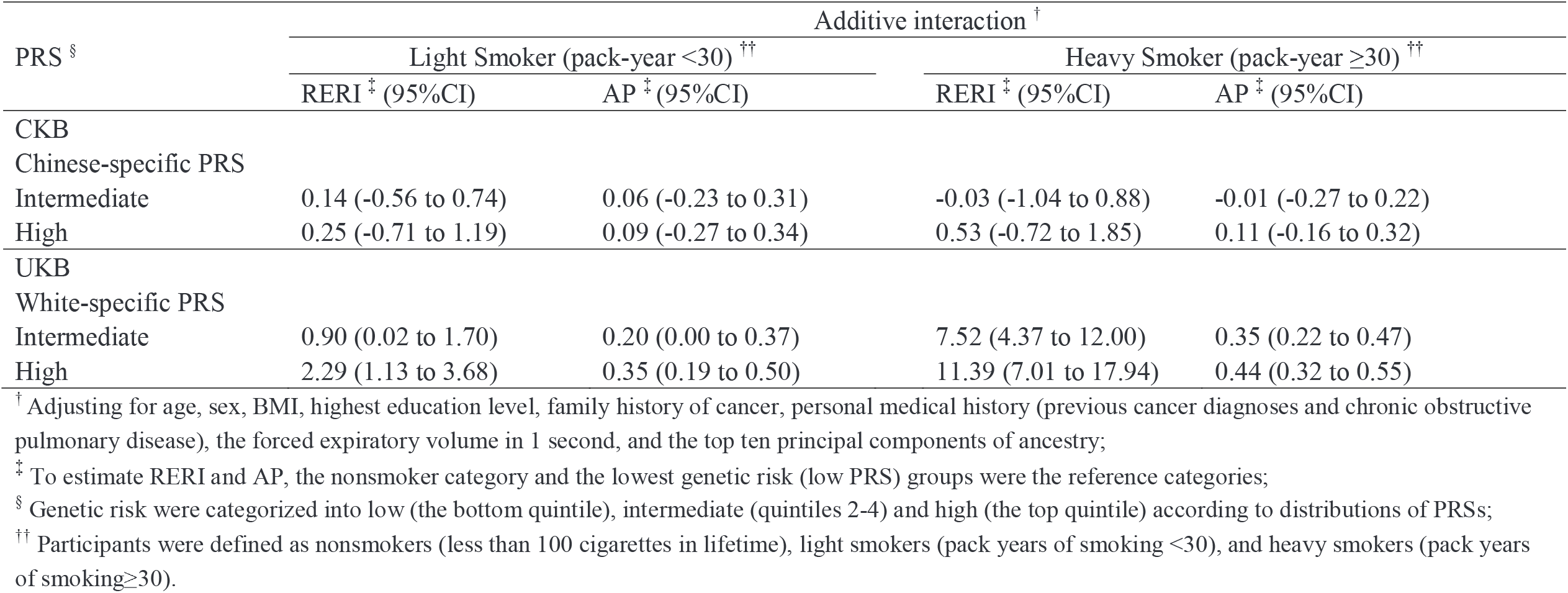
Interactions between genetic risk and pack-years of smoking on the risk of incident lung cancer in the CKB and the UKB

| PRS <sup>§</sup> | Additive interaction <sup>†</sup> |  |  |  |
| --- | --- | --- | --- | --- |
|  | Light Smoker (pack-year <30) <sup>††</sup> |  | Heavy Smoker (pack-year ≥30) <sup>††</sup> |  |
|  | RERI <sup>‡</sup> (95%CI) | AP <sup>‡</sup> (95%CI) | RERI <sup>‡</sup> (95%CI) | AP <sup>‡</sup> (95%CI) |
| CKB |  |  |  |  |
| Chinese-specific PRS |  |  |  |  |
| Intermediate | 0.14 (-0.56 to 0.74) | 0.06 (-0.23 to 0.31) | -0.03 (-1.04 to 0.88) | -0.01 (-0.27 to 0.22) |
| High | 0.25 (-0.71 to 1.19) | 0.09 (-0.27 to 0.34) | 0.53 (-0.72 to 1.85) | 0.11 (-0.16 to 0.32) |
| UKB |  |  |  |  |
| White-specific PRS |  |  |  |  |
| Intermediate | 0.90 (0.02 to 1.70) | 0.20 (0.00 to 0.37) | 7.52 (4.37 to 12.00) | 0.35 (0.22 to 0.47) |
| High | 2.29 (1.13 to 3.68) | 0.35 (0.19 to 0.50) | 11.39 (7.01 to 17.94) | 0.44 (0.32 to 0.55) |

<sup>†</sup> Adjusting for age, sex, BMI, highest education level, family history of cancer, personal medical history (previous cancer diagnoses and chronic obstructive pulmonary disease), the forced expiratory volume in 1 second, and the top ten principal components of ancestry;
<sup>‡</sup> To estimate RERI and AP, the nonsmoker category and the lowest genetic risk (low PRS) groups were the reference categories;
<sup>§</sup> Genetic risk were categorized into low (the bottom quintile), intermediate (quintiles 2-4) and high (the top quintile) according to distributions of PRSs;
<sup>††</sup> Participants were defined as nonsmokers (less than 100 cigarettes in lifetime), light smokers (pack years of smoking <30), and heavy smokers (pack years of smoking ≥30).

**Figure 3.**
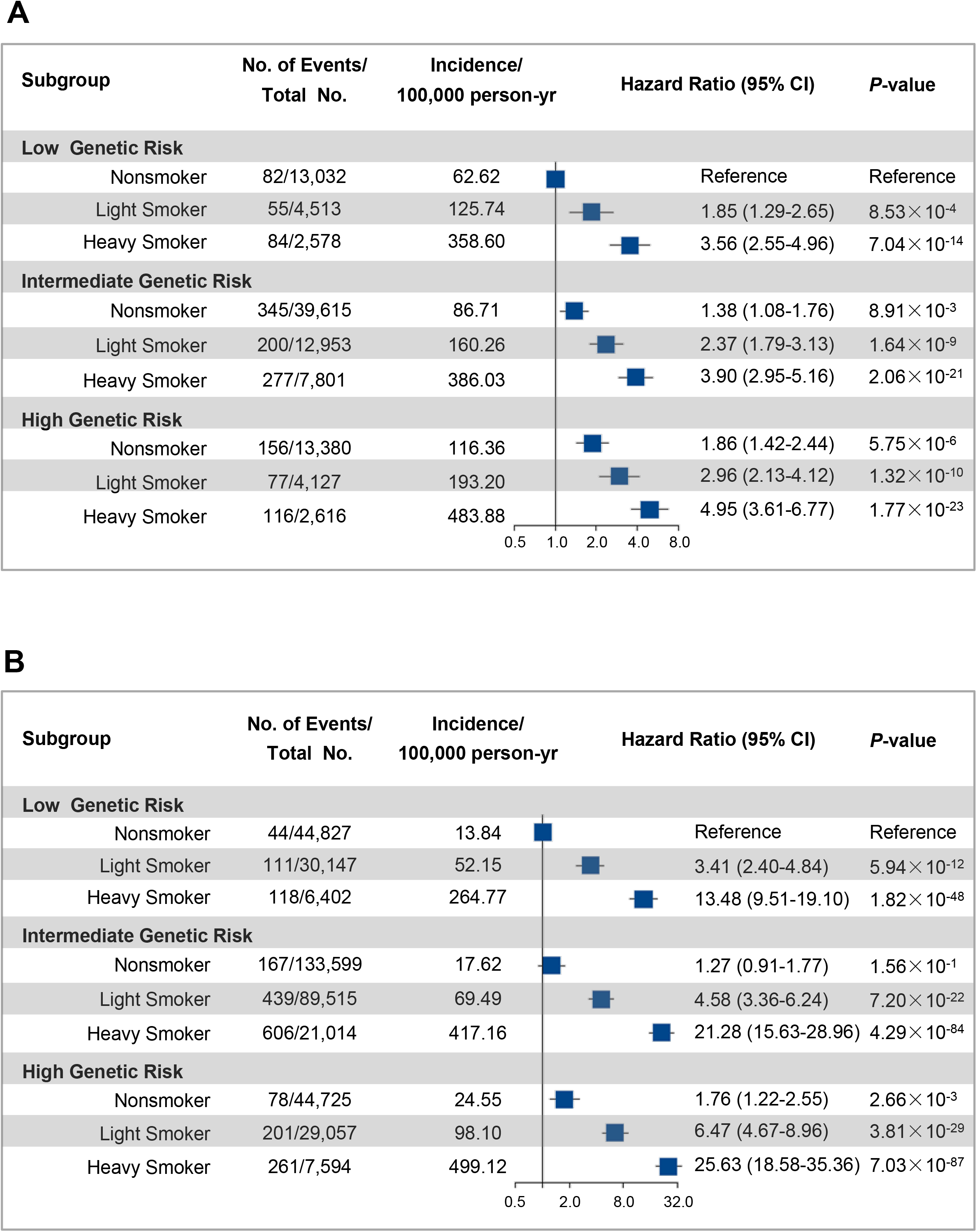
Risk of incident lung cancer according to ethnic-specific polygenic risk scores (PRSs) and pack-years of smoking categories in the CKB (A) and UKB cohorts (B). The hazard ratios were estimated using Cox proportional-hazard models with adjustment for age, sex, BMI, highest education level, family history of cancer, personal medical history, the forced expiratory volume in 1 second, and the top ten principal components of ancestry.

A higher standardized 5-year absolute risk of lung cancer was observed in nonsmokers of the CKB than that in the UKB (3.07 *versus* 0.66 per 1000 person-years), whereas similar absolute risks were observed in heavy smokers between two cohorts (10.19 *versus* 11.60 per 1000 person-years) (**Figure 4**). As a result, a greater risk reduction (never smokers *versus* heavy smokers) was found in the UKB than that in the CKB (*P*<0.001), with an estimation of 10.95 (95% CI, 9.89-11.96) and 7.12 (95% CI, 5.87-8.37) per 1000 person-years, respectively. In participants of low genetic risk, the reductions were comparable between two cohorts (6.54 [95%CI, 4.69-8.15] *versus* 7.05 [95%CI, 4.38-9.43] per 1000 person-years in the UKB and CKB, respectively). However, in those of high genetic risk, the reductions were expanded in the UKB (14.57 [95%CI, 12.06-17.05] per 1000 person-years) compared with that in the CKB (8.01 [95%CI, 5.27-10.47] per 1000 person-years). Similar patterns were noted by reanalyzing with the trans-ancestry PRS (**Figure E7**) or reclassifying genetic risk levels by quartile or tertiles of the PRS (**Table E17-18**).

**Figure 4.**
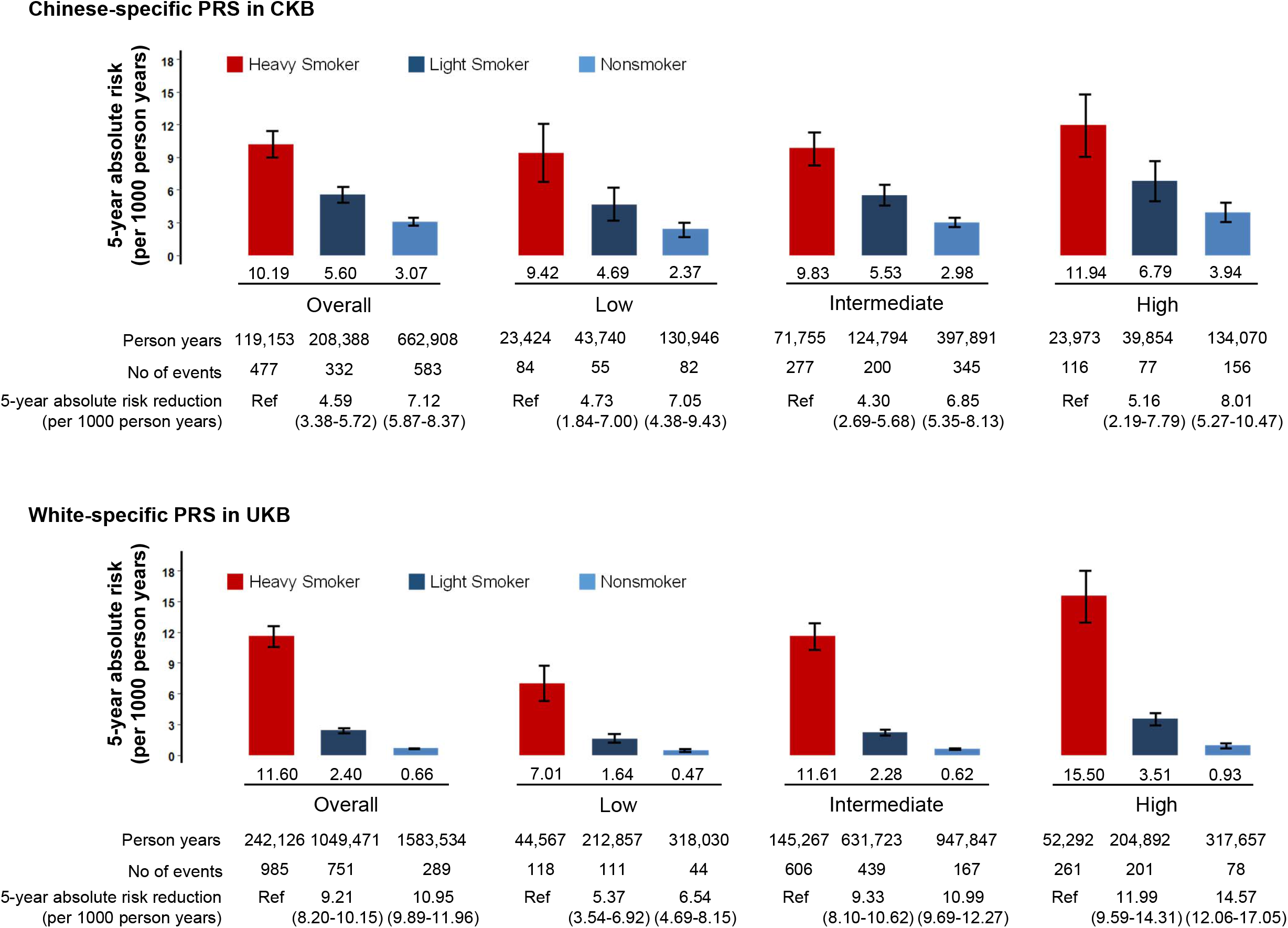
Absolute risk and risk reduction of incident lung cancer according to pack-years of smoking within each genetic risk category defined by ethnic-specific polygenic risk score (PRS). Genetic risk was categorized into low (the bottom quintile), intermediate (quintiles 2-4) and high (the top quintile) according to distributions of PRSs. The 5-year absolute risks were standardized for age according to the mean in CKB and UKB synchronously. The HRs were estimated using Cox proportional hazards regression with adjustment for age, sex, BMI, highest education level, family history of cancer, personal medical history, the forced expiratory volume in 1 second, and the top ten principal components of ancestry. The 5-year absolute risk reduction and 95% CI were generated by drawing 1000 bootstrap samples from the estimation dataset.

## Discussion

In the present study, we integrated the largest GWASs of lung cancer and nationwide prospective cohorts in populations of Chinese and white descents to disaggregate the effects of genetic risk and smoking on lung cancer incidences. Our results indicated that even though the Chinese-specific and White-specific PRSs were consistently associated with risk of lung cancer respectively, ethnic differences were observed for the compositions, relative risks, mediation effects, and additive interactions with smoking of the two PRSs. Therefore, to the best of our knowledge, this study is the first to provide convincing evidences that ethnic differences in genetic background are involved in “the smoking paradox” between Chinese population and white population.

Studies have shown that PRS, as an indicator of genetic risk, can efficiently predict incidence of site-specific cancer and overall cancer (9, 12, 17, 18). For lung cancer, ethnic-specific PRSs have been shown to be effective in discriminating subpopulations at high risk of lung cancer and informing the optimal lung cancer screening strategy (9, 10). Although only 23 SNPs were included in the construction of white-specific PRS, a similar association was observed compared with that of previously used PRS with 114 SNPs in the UKB (10). In consistent with previous findings of ethnic heterogeneity in some lung cancer susceptibility loci (8), the present study provided a comprehensive panorama of genetic differences for lung cancer, and highlighted the role of smoking-related genetic loci in lung cancer susceptibility of white population. Furthermore, our findings also support the notion that an ethnic-specific PRS predicts individual risk more accurately (19).

Several possible explanations have been proposed for the “smoking paradox”, such as difference in epidemics of cigarettes exposure between developed and developing countries (3, 4) or in toxicity and filters changing over time in different countries (20). Our study further indicated that interactions between genetic risk and smoking may be another one of the important reasons for the “smoking paradox” in white population, in that a relative excess risk of up to 44% could be explained by the observed additive interaction. The interactions may be due to smoking addiction-related susceptibility loci (such as *CHRNA5* in 15q25.1) have higher frequency and stronger effects in white population than in Chinese population. However, the interaction was unlikely to be simply mediated by the number of cigarettes smoked, because only 2.06% of the associations between PRS and lung cancer risk could be explained by smoking amount. These results suggested that high genetic risk was probably associated with multiple risk mechanisms for lung cancer in white population, including delayed smoking cessation, increased intensity of smoking exposure, and potential impact on treatment response (21). These findings reveal the genetic basis of the strong association between smoking and lung cancer in white population, and could help explain the huge reduced lung cancer burden in white population after tobacco control during the past decades (22).

Another possible explanation for the “smoking paradox” is a high incidence of lung cancer in nonsmokers of East Asian (23), which is also supported by our cohort study. For example, we found that the estimated age-standardized 5-year absolute risks in nonsmokers were more than 3 times higher in the CKB than that in the UKB across genetic risk groups. In addition, differences in genetic risk are probably not the reason for the observed high incidence of lung cancer in nonsmokers of Chinese, because we also observed a lower distribution of trans-ancestry PRS in the CKB than that in the UKB (**Figure E8**). These results indicated that environmental risk factors beyond smoking, especially those of high exposure levels in Chinese populations, need to be further explored in relation to this discordance. For example, recent studies have shown that exposure to high concentrations of PM_2.5_ in the ambient environment could increase lung cancer risk in both Chinese population and white population; however, the average exposure concentration was estimated to be 65 μg/m^3^ in China between 2000-2015, compared with 10 μg/m^3^ in the UK around 2010 (24-26). Therefore, our results indicate that further efforts are needed to clarify and control the causes for the high incidence of lung cancer in nonsmokers of Chinese population.

Here, we further showed that the absolute risks of lung cancer were reduced for nonsmokers compared with smokers across genetic risk groups in Chinese population and white population, consistently. Therefore, our findings support the notion that public efforts to promote smoking cessation will lead to an overall reduction of lung cancer risk across ethnic groups (27). However, we observed that the benefits of tobacco control would be greater in white population than that in Chinese population, especially for those at high genetic risk. This indicated that precision interventions for smoking cessation based on genetic risk are feasible for white population (28), but not for Chinese population. Taken together, our results showed that it was more complicated to control lung cancer epidemic in China, and comprehensive policies against smoking and nonsmoking risk factors should be made to lower its high incidence.

The present study has several strengths, including the large sample size from two well-ethnically defined lung cancer GWASs and two well established nationwide prospective cohorts in China and the UK; the standardized approaches to assess individual genetic risk of lung cancer simultaneously; and a series of sensitivity analyses to show the robustness of the findings. Nevertheless, we also acknowledge several limitations. First, the sample size of lung cancer GWAS is obviously larger in white population than that in Chinese, which may lead to the reported lung cancer susceptibility loci more relevant to white population. Second, information on smoking was mainly self-reported and only measured once; thus, misclassification was inevitable and behavioral changes during the follow-up may have an effect on risk estimates. Third, the calculation of pack-years assumed that cumulative number of cigarettes had the same health effects, which might not be true. Fourth, although personal characteristics and comorbidities were controlled in the present study, additional potential confounders, e.g. occupational exposure were not assessed in the cohorts, might result in potential residual confounding. Finally, both CKB and UKB were not designed to include a representative study population (11, 13); therefore, further investigations are warranted to evaluate to what degree these findings may be generalized to the general population.

In summary, our comprehensive analysis demonstrated that ethnic differences of genetic factors were involved in “the smoking paradox” observed between Chinese population and white population. Specifically, our results highlighted that white population were more susceptible to lung cancer caused by smoking and had a greater benefits of smoking cessation, especially in high genetic risk population. Moreover, nonsmokers of Chinese had consistently higher absolute risk of lung cancer than those of white population across genetic risk groups. These results collectively indicate that tobacco control alone is not enough to reduce the burden of lung cancer in China, and more comprehensive policies against smoking and nonsmoking risk factors should be made to lower the high incidence of lung cancer in China.

## Data Availability

Researchers can apply to use the UK Biobank and the China Kadoorie Biobank resource and access the data used. No additional data are available.

## Abbreviations

AP: the attributable proportion because of the interaction
CI: confidence interval
CKB: the China Kadoorie Biobank
GWAS: genome-wide association study
HR: hazards ratio
ICD-10: the 10th Revision of International Classification of Diseases
OR: odds ratio
PRS: polygenic risk score
RERI: relative excess risk due to interaction
RR: relative risk
SNP: single-nucleotide polymorphism
UKB: the UK Biobank

## Supplementary Information

The online version contains supplementary material.

## Acknowledgements

This research was conducted using the UK Biobank (Application Number: 48700) and China Kadoorie Biobank. We thank the investigators and participants involved in the UK Biobank and China Kadoorie Biobank for their contributions to this study. We also thank to all principal investigators who participated in the INTEGRAL-ILCCO OncoArray Project.

## Authors’ Contributions

HS, LL and ZH supervised the entire project and design the work. MZ, JL, YH, and HM contributed to the data analysis, data interpretation, and writing of the report. XW, MJ, ZM, CW, JD, NL, YG, RW, IM, RH, DC, CY, GJ, ZC, QW, and CA contributed to the discussion, data interpretation and revised the manuscript. All authors reviewed or revised the manuscript and approved the final draft for submission. The authors declare no conflicts of interest.

## Declarations

### Ethics approval and consent to participate

The CKB has been approved by the Ethical Review Committee of the Oxford Tropical Research Ethics Committee, University of Oxford and the Chinese Center for Disease Control and Prevention. The UKB has been approved by the multicenter Research Ethics Committee, the National Information Governance Board for Health and Social Care in England and Wales, and the Community Health Index Advisory Group in Scotland.

### Consent for publication

Not applicable.

