## Supplementary material for "Ethnic differences of genetic risk and smoking in lung cancer: two prospective cohort studies": Table E1-18,Figure E1-8

**Contents**

Supplementary Methods

Quality control and PRS construction

Genotyping and imputation

Assessment of covariates

Statistical analyses

References

Table E1. Incidence of lung cancer in different subgroups of the CKB and the UKB.

Table E2. Association results of the 81 reported lung cancer susceptibility SNPs in each GWAS dataset.

Table E3. Associations of cross-used ethnic-specific polygenic risk score (PRS) with incident lung cancer in the CKB and the UKB.

Table E4. Risk of incident lung cancer additionally adjusting for environmental exposures.

Table E5. Risk of incident lung cancer according to different risk levels of polygenic risk score (PRS).

Table E6. Risk of incident lung cancer after excluding participants with missing covariates.

Table E7. Risk of incident lung cancer after excluding incident cases during the first year of follow-up.

Table E8. Risk of incident lung cancer according to levels of pack-years of smoking in the CKB and the UKB.

Table E9. Risk of incident lung cancer according to smoking status in the CKB and the UKB.

Table E10. Associations between polygenic risk scores (PRSs) and pack-years of smoking.

Table E11. Mediation analysis of pack-years of smoking on the associations between polygenic risk scores (PRSs) and lung cancer.

Table E12. Interactions between trans-ancestry polygenic risk scores (PRSs) and pack-years of smoking on the risk of incident lung cancer in the CKB and the UKB.

Table E13. Interactions between genetic risk and pack-years of smoking on the risk of incident lung cancer additionally adjusting for environmental exposures.

Table E14. Interactions between genetic risk and pack-years of smoking on the risk of incident lung cancer according to different risk levels of polygenic risk scores (PRSs) in the CKB and the UKB.

Table E15. Interactions between genetic risk and pack-years of smoking on the risk of incident lung cancer after excluding incident cases during the first year of follow-up.

Table E16. Interactions between genetic risk and pack-years of smoking on the risk of incident lung cancer after excluding participants with missing covariates.

Table E17. Risk of incident cancer according to pack-years of smoking within each genetic risk (quartiles) level in the CKB and the UKB.

Table E18. Risk of incident cancer according to pack-years of smoking within each genetic risk (tertiles) level in the CKB and the UKB.

Figure E1. Study design and workflow.

Figure E2. Flowchart for the calculation of polygenic risk scores (PRSs).

Figure E3. The relationship of ethnic-specific polygenic risk scores (PRSs) with incident lung cancer in CKB and UKB cohorts.

Figure E4. Mediation analysis of smoking on associations between PRS and incident lung cancer risk in the CKB (A) and UKB cohorts (B).

Figure E5. Flowchart for the calculation of trans-ancestry polygenic risk scores (PRSs).

Figure E6. Risk of incident lung cancer according to trans-ancestry polygenic risk scores (PRSs) and pack-years of smoking categories in the CKB (A) and UKB cohorts (B).

Figure E7. Absolute risk and risk reduction of incident lung cancer according to pack-years of smoking within each genetic risk category defined by trans-ancestry PRS.

Figure E8. The distributions of trans-ancestry polygenic risk scores (PRSs) in the CKB and the UKB.

**Supplementary Methods**

**Quality control and PRS construction**

To ensure the efficiency of the PRS, we applied several criteria to exclude the redundant variants, i.e., (i) variants with a minor allele frequency less than 0.5% in corresponding ethnicity; (ii) SNPs in linkage disequilibrium (LD, r^2^≥0.1); (iii) variants with p-value > 0.05 to be excluded after multiple comparisons; and (iv) the SNP with the strongest association were kept in each LD block. The candidate SNPs were selected for Chinese and white population, respectively, based on the GWAS association results (Table E1) and LD information from 1000 Genomes project. The SNP selection process was performed with “--clump” in PLINK v1.90b3b. At last, a total of 19 SNPs and 23 SNPs were retained for the calculation of Chinese-specific (PRS-19) and White-specific PRSs (PRS-23), respectively. Although the SNPs were independent within PRS-19 and PRS-23, respectively, we found four SNPs in PRS-19 and PRS-23 were in high LD (r^2^>0.8), which were mainly due to the differences of the most significant SNPs of the same loci in different datasets. To simplify the comparison between different populations, we replaced the candidate SNPs in PRS-23 with their high LD SNPs in PRS-19, including rs11375254 for rs13080835 (r^2^=0.89), rs401681 for rs465498 (r^2^=0.96), rs10429489 for rs885518 (r^2^=0.84), and rs55768116 for rs1056562 (r^2^=0.84). The detail information of PRS-19 and PRS-23 was shown in Figure E2.

**Genotyping and imputation**

The genotyping process and DNA arrays used in the CKB and UKB cohorts have been described elsewhere in more detail (1, 2). Briefly, participants in CKB were genotyped using a custom-designed Affymetrix Axiom array (optimized for Chinese participants), which included approximately 700,000 markers. The genotype data were imputed with SHAPEIT2 and IMPUTE2 by using the 1000 Genomes phase3 as the reference (3, 4). Participants in the UKB were genotyped using Applied Biosystems™ UK BiLEVE Axiom™ Array (49950 participants) or Applied Biosystems™ UK Biobank Axiom Array (438427 participants) by Affymetrix, which consisted of more than 800,000 markers (2). Imputation was performed with SHAPEIT3 and IMPUTE3 based on merged panels of UK10K and 1000 Genomes phase3 (3, 4).

**Assessment of covariates**

Covariates were obtained through questionnaires, including age, sex, education level, family history of cancer, and personal medical history. Body weight and height were measured at baseline, with BMI calculated as weight (kg)/ (height (m)^2^). Spirometry was performed using a portable handheld ‘Micro spirometer’ (Micro Medical Limited, CKB) or a handheld pneumotachograph spirometer (Pneumotrac 6800, UKB) in accordance with modified American Thoracic Society (ATS) procedures. Missing data in continuous covariates were imputed with the conditional average of each respective variable by sex, and missing values of categorical covariates were classified as “unknown” category for each corresponding variable.

**Statistical analyses**

We assessed lung cancer risk in participants from the enrolment until the time of lung cancer diagnosis, death, or the end of follow-up, whichever occurred first. We used linear regression models to assess associations between the PY smoked and individual PRS. We also assessed a potential nonlinear relationship between the PRS and lung cancer risk by use of restricted cubic spline analysis. Cox proportional hazard models were used to assess associations between genetic factors and smoking with lung cancer incidence and to estimate hazards ratios (HRs) and 95% confidence intervals (CIs) with adjustment for age, sex, smoking status, BMI, highest education level, family history of cancer, personal medical history (previous cancer diagnoses and chronic obstructive pulmonary disease), forced expiratory volume in 1 second, and the top ten principal components of ancestry (5). Schoenfeld residuals were used to test the proportional hazards assumption. The genetic risk was categorized into low (the bottom quintile), intermediate (quintiles 2-4) and high (the top quintile) based on distributions of PRSs, as described previously (6, 7). We compared the incident lung cancer risk for participants at high or intermediate genetic risk with those at low genetic risk using Cox regression analysis. Similarly, we compared the effect of heavy or light smoking on risk of incident lung cancer to that of nonsmokers. Relative excess risk due to interaction (RERI) and the attributable proportion because of the interaction (AP) were calculated to measure the interaction on the additive scale based on coefficients of the product term (8). The 95% CIs of the RERI and AP were derived by drawing 5000 bootstrap samples from the estimation dataset, which would include 0 if there were no additive interactions (9). The mediation proportion by the mediator was calculated by comparing estimates from models with and without the hypothesized mediator (10). We then calculated cumulative risk as the incidence of lung cancer occurring in a given group during follow-up. We also calculated absolute risk reduction as the difference in lung cancer incidence between given groups over a 5-year period, and the 95% CIs for the absolute risk reduction were generated by drawing 1000 bootstrap samples from the estimation dataset.

We performed several sensitivity analyses to examine the robustness of the results: (1) trans-ancestry PRSs were generated simultaneously to define genetic risk of the participants; (2) additional environmental exposures were included in the model, including passive smoking and ambient PM_2.5_ concentration; (3) genetic risk levels were reclassified by quartile or tertiles; (4) smoking status was reclassified as never, former, and current smokers; (5) participants who were diagnosed with lung cancer within the first year of follow-up were excluded; and (6) analysis were restricted to participants with complete covariate data for comparison with the results of imputation. All P-values were two-sided, and P<0.05 was considered statistically significant. All statistical analyses were performed in R (version 3.5).

Table E1. Incidence of lung cancer in different subgroups of CKB and UKB

| Age group | Lung cancer incidence in CKB (per 100,000 person-year) ^†^ | | | | | | | |  | Lung cancer incidence in UKB (per 100,000 person-year) ^†^ | | | | | | | |
| --- | --- | --- | --- | --- | --- | --- | --- | --- | --- | --- | --- | --- | --- | --- | --- | --- | --- |
|  | Nonsmoker | |  | Light smoker | |  | Heavy smoker | |  | Nonsmoker | |  | Light smoker | |  | Heavy smoker | |
|  | Male | Female |  | Male | Female |  | Male | Female |  | Male | Female |  | Male | Female |  | Male | Female |
| -45 | 23.84 | 21.64 |  | 30.58 | 33.83 |  | 94.50 | - |  | 2.12 | 4.27 |  | 8.61 | 12.12 |  | 37.93 | 188.02 |
| 45-50 | 40.07 | 45.57 |  | 77.51 | 53.35 |  | 121.06 | - |  | 4.06 | 6.89 |  | 18.08 | 23.54 |  | 131.50 | 144.52 |
| 50-55 | 65.54 | 68.41 |  | 122.04 | 111.50 |  | 236.17 | 262.91 |  | 9.38 | 7.84 |  | 48.11 | 31.30 |  | 206.99 | 302.50 |
| 55-60 | 101.00 | 89.11 |  | 154.59 | 187.18 |  | 339.81 | 225.83 |  | 11.30 | 24.32 |  | 62.17 | 67.06 |  | 334.08 | 409.30 |
| 60-65 | 160.25 | 142.83 |  | 252.57 | 284.71 |  | 442.99 | 617.12 |  | 25.41 | 49.00 |  | 235.02 | 191.24 |  | 1,902.44 | 1,786.82 |
| 65-70 | 200.44 | 177.45 |  | 371.15 | 522.71 |  | 650.30 | 599.71 |  | 39.65 | 36.23 |  | 154.49 | 158.81 |  | 606.49 | 679.82 |
| 70- | 227.78 | 182.89 |  | 480.61 | 505.73 |  | 810.58 | 777.27 |  | - | - |  | - | - |  | - | - |

^†^ Participants were defined as nonsmokers (less than 100 cigarettes in lifetime), light smokers (pack years of smoking, PY<30), and heavy smokers (PY≥30);

Table E2. Association results of the 81 reported lung cancer susceptibility SNPs in each GWAS dataset

| SNP | cytoBand | Pos | Allele | |  | Chinese population | | | | |  | White population | | | | |  | Trans-ancestry Meta | | | | |
| --- | --- | --- | --- | --- | --- | --- | --- | --- | --- | --- | --- | --- | --- | --- | --- | --- | --- | --- | --- | --- | --- | --- |
|  |  |  | Effect | Alt |  | EAF^†^ | OR | L95 | U95 | *P* |  | EAF^†^ | OR | L95 | U95 | *P* |  | EAF^†^ | OR | L95 | U95 | *P* |
| rs71658797 | 1p31.1 | 77967507 | A | T |  | 0.00 | NA | NA | NA | NA |  | 0.10 | 1.14 | 1.09 | 1.18 | 3.25×10^-11^ |  | NA | NA | NA | NA | NA |
| rs17038564 | 2p14 | 65496058 | G | A |  | 0.19 | 1.11 | 1.05 | 1.16 | 5.45×10^-5^ |  | 0.04 | 1.06 | 0.99 | 1.13 | 8.39×10^-2^ |  | 0.16 | 1.10 | 1.06 | 1.15 | 1.13×10^-5^ |
| rs3769821 | 2q33.1 | 202123430 | C | T |  | 0.29 | 1.06 | 1.01 | 1.10 | 1.08×10^-2^ |  | 0.34 | 1.05 | 1.02 | 1.07 | 2.17×10^-4^ |  | 0.32 | 1.08 | 1.05 | 1.11 | 4.45×10^-8^ |
| rs2293607 | 3q26.2 | 169482335 | T | C |  | 0.45 | 1.12 | 1.08 | 1.17 | 2.54×10^-9^ |  | 0.77 | 1.04 | 1.02 | 1.07 | 2.29×10^-3^ |  | 0.60 | 1.10 | 1.06 | 1.13 | 1.82×10^-10^ |
| rs11375254 | 3q28 | 189343242 | T | TA |  | 0.51 | 1.21 | 1.16 | 1.26 | 2.92×10^-21^ |  | 0.54 | 1.08 | 1.04 | 1.11 | 4.19×10^-5^ |  | 0.52 | 1.13 | 1.10 | 1.16 | 1.35×10^-20^ |
| rs4488809 | 3q28 | 189356261 | T | C |  | 0.48 | 1.20 | 1.15 | 1.25 | 2.72×10^-20^ |  | 0.51 | 1.06 | 1.03 | 1.08 | 1.43×10^-6^ |  | 0.50 | 1.13 | 1.10 | 1.16 | 1.03×10^-20^ |
| rs13080835 | 3q28 | 189357199 | G | T |  | 0.48 | 1.20 | 1.15 | 1.25 | 2.77×10^-20^ |  | 0.51 | 1.06 | 1.03 | 1.08 | 1.25×10^-6^ |  | 0.50 | 1.13 | 1.10 | 1.16 | 9.93×10^-21^ |
| rs13314271 | 3q28 | 189357602 | T | C |  | 0.48 | 1.20 | 1.15 | 1.25 | 2.99×10^-20^ |  | 0.51 | 1.06 | 1.03 | 1.08 | 1.26×10^-6^ |  | 0.50 | 1.13 | 1.10 | 1.16 | 1.00×10^-20^ |
| rs10937405 | 3q28 | 189383183 | C | T |  | 0.70 | 1.16 | 1.11 | 1.21 | 1.29×10^-11^ |  | 0.55 | 1.04 | 1.02 | 1.06 | 1.25×10^-3^ |  | 0.63 | 1.09 | 1.06 | 1.12 | 2.47×10^-10^ |
| rs2131877 | 3q29 | 194858374 | A | G |  | 0.56 | 1.03 | 0.99 | 1.07 | 1.32×10^-1^ |  | 0.18 | 1.01 | 0.98 | 1.04 | 4.14×10^-1^ |  | 0.40 | 1.03 | 1.00 | 1.07 | 2.26×10^-2^ |
| rs13167280 | 5p15 | 1280477 | A | G |  | 0.21 | 1.30 | 1.24 | 1.37 | 2.33×10^-25^ |  | 0.14 | 1.12 | 1.08 | 1.16 | 8.14×10^-9^ |  | 0.17 | 1.24 | 1.19 | 1.28 | 4.59×10^-32^ |
| rs7705526 | 5p15 | 1285974 | A | C |  | 0.41 | 1.31 | 1.26 | 1.36 | 3.91×10^-41^ |  | 0.34 | 1.12 | 1.10 | 1.15 | 1.01×10^-18^ |  | 0.37 | 1.20 | 1.17 | 1.24 | 1.06×10^-42^ |
| rs2736100 | 5p15 | 1286516 | C | A |  | 0.42 | 1.29 | 1.24 | 1.34 | 8.92×10^-38^ |  | 0.52 | 1.09 | 1.05 | 1.12 | 1.15×10^-6^ |  | 0.48 | 1.17 | 1.14 | 1.20 | 1.36×10^-33^ |
| rs2853677 | 5p15 | 1287194 | G | A |  | 0.39 | 1.30 | 1.25 | 1.35 | 1.34×10^-38^ |  | 0.42 | 1.12 | 1.09 | 1.15 | 2.66×10^-18^ |  | 0.42 | 1.20 | 1.17 | 1.23 | 3.27×10^-41^ |
| rs4635969 | 5p15 | 1308552 | G | A |  | 0.87 | 1.24 | 1.16 | 1.31 | 2.18×10^-11^ |  | 0.81 | 1.16 | 1.12 | 1.19 | 1.17×10^-21^ |  | 0.83 | 1.19 | 1.14 | 1.23 | 1.96×10^-20^ |
| rs4975616 | 5p15 | 1315660 | A | G |  | 0.82 | 1.18 | 1.12 | 1.24 | 4.26×10^-10^ |  | 0.59 | 1.14 | 1.12 | 1.17 | 2.99×10^-29^ |  | 0.65 | 1.18 | 1.15 | 1.22 | 3.60×10^-29^ |
| rs401681 | 5p15 | 1322087 | C | T |  | 0.68 | 1.16 | 1.11 | 1.20 | 8.21×10^-12^ |  | 0.57 | 1.15 | 1.12 | 1.17 | 3.25×10^-30^ |  | 0.61 | 1.17 | 1.14 | 1.20 | 7.37×10^-30^ |
| rs465498 | 5p15 | 1325803 | A | G |  | 0.82 | 1.19 | 1.13 | 1.26 | 4.24×10^-11^ |  | 0.58 | 1.15 | 1.12 | 1.18 | 2.68×10^-32^ |  | 0.64 | 1.18 | 1.15 | 1.22 | 1.79×10^-30^ |
| rs31489 | 5p15 | 1342714 | C | A |  | 0.82 | 1.19 | 1.13 | 1.25 | 1.07×10^-10^ |  | 0.60 | 1.15 | 1.12 | 1.17 | 1.02×10^-29^ |  | 0.66 | 1.17 | 1.14 | 1.21 | 1.30×10^-26^ |
| rs2895680 | 5q32 | 146644115 | C | T |  | 0.30 | 1.05 | 1.01 | 1.09 | 2.69×10^- 2^ |  | 0.28 | 1.01 | 0.98 | 1.03 | 5.68×10^-1^ |  | 0.27 | 1.03 | 1.01 | 1.07 | 2.09×10^-2^ |
| rs7741164 | 6p21.1 | 41493412 | A | G |  | 0.34 | 1.16 | 1.11 | 1.21 | 3.71×10^-12^ |  | 0.03 | 0.97 | 0.90 | 1.06 | 5.50×10^-1^ |  | 0.30 | 1.13 | 1.09 | 1.18 | 8.07×10^-10^ |
| rs1853837 | 6p21.1 | 41497035 | A | C |  | 0.34 | 1.15 | 1.10 | 1.20 | 5.48×10^-11^ |  | 0.11 | 1.01 | 0.97 | 1.05 | 6.77×10^-1^ |  | 0.26 | 1.12 | 1.08 | 1.15 | 1.21×10^-10^ |
| rs115790694 | 6p21.3 | 29875992 | A | G |  | 0.04 | 1.20 | 1.07 | 1.34 | 1.46×10^-3^ |  | 0.18 | 1.14 | 1.10 | 1.18 | 2.23×10^-14^ |  | 0.15 | 1.16 | 1.11 | 1.21 | 4.60×10^-11^ |
| rs2523571 | 6p21.3 | 31329691 | A | T |  | 0.04 | 1.02 | 0.92 | 1.14 | 7.23×10^-1^ |  | 0.11 | 1.17 | 1.13 | 1.21 | 1.09×10^-16^ |  | 0.08 | 1.11 | 1.05 | 1.16 | 7.49×10^-5^ |
| rs3094604 | 6p21.3 | 31434111 | G | A |  | 0.17 | 0.96 | 0.92 | 1.02 | 1.93×10^-1^ |  | 0.16 | 1.15 | 1.12 | 1.19 | 5.29×10^-19^ |  | 0.17 | 1.05 | 1.01 | 1.08 | 1.04×10^-2^ |
| rs9469031 | 6p21.3 | 31595795 | C | T |  | 0.98 | 1.12 | 0.96 | 1.30 | 1.43×10^-1^ |  | NA | NA | NA | NA | NA |  | NA | NA | NA | NA | NA |
| rs3817963 | 6p21.3 | 32368087 | C | T |  | 0.25 | 1.06 | 1.02 | 1.11 | 7.16×10^-3^ |  | 0.27 | 0.99 | 0.97 | 1.02 | 5.52×10^-1^ |  | 0.26 | 1.02 | 0.99 | 1.05 | 2.53×10^-1^ |
| rs2395185 | 6p21.3 | 32433167 | T | G |  | 0.37 | 1.10 | 1.06 | 1.15 | 2.54×10^-6^ |  | 0.30 | 0.98 | 0.96 | 1.01 | 1.70×10^-1^ |  | 0.33 | 1.05 | 1.02 | 1.08 | 1.12×10^-3^ |
| rs3115672 | 6p21.33 | 31727897 | T | C |  | 0.00 | NA | NA | NA | NA |  | 0.10 | 1.18 | 1.14 | 1.23 | 3.47×10^-18^ |  | NA | NA | NA | NA | NA |
| rs200847762 | 6p21.33 | 32097148 | A | G |  | 0.00 | NA | NA | NA | NA |  | 0.00 | NA | NA | NA | NA |  | NA | NA | NA | NA | NA |
| rs2523546 | 6p21.33 | 31332920 | T | C |  | 0.01 | NA | NA | NA | NA |  | 0.11 | 1.17 | 1.13 | 1.21 | 4.44×10^-17^ |  | NA | NA | NA | NA | NA |
| rs1264308 | 6p21.33 | 30879987 | A | G |  | 0.01 | NA | NA | NA | NA |  | 0.11 | 1.18 | 1.14 | 1.22 | 9.20×10^-19^ |  | NA | NA | NA | NA | NA |
| rs3117582 | 6p21.33 | 31620520 | C | A |  | 0.00 | NA | NA | NA | NA |  | 0.10 | 1.18 | 1.14 | 1.23 | 9.48×10^-19^ |  | NA | NA | NA | NA | NA |
| rs5879422 | 6q22.1 | 117784658 | T | TTG |  | 0.51 | 1.09 | 1.05 | 1.13 | 1.75×10^-5^ |  | 0.51 | 1.07 | 1.04 | 1.11 | 3.20×10^-5^ |  | 0.51 | 1.08 | 1.05 | 1.11 | 4.36×10^-9^ |
| rs9387478 | 6q22.2 | 117786180 | C | A |  | 0.50 | 1.09 | 1.05 | 1.13 | 2.07×10^-5^ |  | 0.51 | 1.03 | 1.01 | 1.06 | 6.14×10^-3^ |  | 0.51 | 1.08 | 1.05 | 1.11 | 8.16×10^-9^ |
| rs6920364 | 6q27 | 167376466 | C | G |  | 0.70 | 1.04 | 1.00 | 1.09 | 4.88×10^-2^ |  | 0.46 | 1.07 | 1.05 | 1.10 | 1.29×10^-8^ |  | 0.56 | 1.06 | 1.03 | 1.09 | 2.61×10^-5^ |
| rs2285947 | 7p15.3 | 21584088 | A | G |  | 0.27 | 1.02 | 0.98 | 1.07 | 2.93×10^-1^ |  | 0.48 | 1.01 | 0.99 | 1.04 | 2.53×10^-1^ |  | 0.39 | 1.02 | 0.99 | 1.05 | 1.85×10^-1^ |
| rs4236709 | 8p12 | 32410110 | G | A |  | 0.18 | 1.17 | 1.11 | 1.23 | 1.30×10^-9^ |  | 0.22 | 1.07 | 1.04 | 1.10 | 5.88×10^-6^ |  | 0.20 | 1.12 | 1.09 | 1.16 | 1.11×10^-12^ |
| rs11780471 | 8p21.2 | 27344719 | G | A |  | 0.00 | NA | NA | NA | NA |  | 0.94 | 1.15 | 1.10 | 1.21 | 1.69×10^-8^ |  | NA | NA | NA | NA | NA |
| rs35201538 | 9p13.3 | 33422488 | C | CT |  | 0.65 | 1.10 | 1.06 | 1.15 | 5.36×10^-6^ |  | 0.71 | 1.03 | 1.00 | 1.07 | 7.64×10^-2^ |  | 0.68 | 1.06 | 1.03 | 1.09 | 1.28×10^-5^ |
| rs10429489 | 9p21.3 | 21787521 | A | G |  | 0.25 | 1.11 | 1.06 | 1.16 | 7.53×10^-6^ |  | 0.10 | 1.09 | 1.05 | 1.13 | 7.08×10^-6^ |  | 0.20 | 1.11 | 1.08 | 1.15 | 6.92×10^-10^ |
| rs885518 | 9p21.3 | 21830157 | G | A |  | 0.26 | 1.09 | 1.05 | 1.14 | 7.62×10^-5^ |  | 0.10 | 1.09 | 1.05 | 1.13 | 2.13×10^-6^ |  | 0.20 | 1.11 | 1.07 | 1.15 | 1.86×10^-9^ |
| rs1333040 | 9p21.3 | 22083404 | C | T |  | 0.31 | 1.03 | 0.99 | 1.08 | 1.40×10^-1^ |  | 0.45 | 1.04 | 1.02 | 1.07 | 2.16×10^-4^ |  | 0.37 | 1.03 | 1.00 | 1.06 | 2.63×10^-2^ |
| rs72658409 | 9p21.3 | 22160087 | C | T |  | 0.93 | 1.10 | 1.02 | 1.19 | 1.04×10^-2^ |  | 0.93 | 1.05 | 1.01 | 1.10 | 2.75×10^-2^ |  | 0.93 | 1.07 | 1.02 | 1.12 | 8.36×10^-3^ |
| rs4573350 | 9q33.2 | 124955115 | T | C |  | 0.26 | 1.09 | 1.05 | 1.14 | 5.88×10^-5^ |  | 0.67 | 1.03 | 1.00 | 1.05 | 4.70×10^-2^ |  | 0.49 | 1.05 | 1.02 | 1.08 | 7.13×10^-4^ |
| rs1663689 | 10p14 | 9025195 | T | C |  | 0.59 | 1.01 | 0.97 | 1.05 | 7.31×10^-1^ |  | 0.80 | 1.01 | 0.98 | 1.04 | 5.52×10^-1^ |  | 0.68 | 1.01 | 0.98 | 1.04 | 4.73×10^-1^ |
| rs11591710 | 10q24.33 | 105687632 | C | A |  | 0.01 | 1.01 | 0.84 | 1.22 | 9.22×10^-1^ |  | 0.14 | 1.07 | 1.04 | 1.11 | 3.53×10^-5^ |  | 0.13 | 1.08 | 1.03 | 1.14 | 1.03×10^-3^ |
| rs12265047 | 10q25.2 | 114487925 | G | A |  | 0.28 | 1.14 | 1.10 | 1.19 | 6.00×10^-10^ |  | 0.02 | 1.04 | 0.97 | 1.11 | 3.26×10^-1^ |  | 0.24 | 1.13 | 1.09 | 1.17 | 7.50×10^-10^ |
| rs7086803 | 10q25.2 | 114498476 | A | G |  | 0.28 | 1.14 | 1.10 | 1.19 | 3.29×10^-10^ |  | 0.02 | 1.02 | 0.95 | 1.09 | 6.48×10^-1^ |  | 0.24 | 1.12 | 1.08 | 1.17 | 3.59×10^-9^ |
| rs55768116 | 11q23.3 | 118108331 | C | A |  | 0.45 | 1.13 | 1.08 | 1.17 | 2.20×10^-9^ |  | 0.51 | 1.07 | 1.04 | 1.09 | 2.42×10^-8^ |  | 0.49 | 1.10 | 1.07 | 1.13 | 2.23×10^-13^ |
| rs1056562 | 11q23.3 | 118125625 | T | C |  | 0.41 | 1.12 | 1.08 | 1.17 | 3.24×10^-9^ |  | 0.48 | 1.07 | 1.04 | 1.09 | 1.92×10^-8^ |  | 0.45 | 1.10 | 1.07 | 1.13 | 3.11×10^-13^ |
| rs7953330 | 12p13.33 | 998819 | C | G |  | 0.26 | 1.01 | 0.97 | 1.06 | 5.68×10^-1^ |  | 0.31 | 0.92 | 0.89 | 0.94 | 6.10×10^-12^ |  | 0.29 | 0.96 | 0.93 | 0.99 | 5.32×10^-3^ |
| rs3748522 | 12p13.33 | 1058688 | C | A |  | 0.32 | 1.02 | 0.98 | 1.06 | 4.03×10^-1^ |  | 0.52 | 1.07 | 1.05 | 1.10 | 8.19×10^-10^ |  | 0.44 | 1.05 | 1.02 | 1.08 | 2.64×10^-4^ |
| rs6489769 | 12p13.33 | 1072965 | T | C |  | 0.53 | 1.02 | 0.98 | 1.06 | 2.85×10^-1^ |  | 0.63 | 1.06 | 1.04 | 1.09 | 8.79×10^-7^ |  | 0.58 | 1.04 | 1.02 | 1.07 | 1.55×10^-3^ |
| rs11610143 | 12q13.13 | 52349071 | C | G |  | 0.70 | 1.07 | 1.02 | 1.11 | 3.33×10^-4^ |  | 0.80 | 0.95 | 0.92 | 1.00 | 3.85×10^-2^ |  | 0.75 | 1.01 | 0.98 | 1.04 | 5.13×10^-1^ |
| rs12296850 | 12q23.1 | 100820085 | G | A |  | 0.75 | 0.99 | 0.94 | 1.03 | 5.55×10^-1^ |  | 0.06 | 0.97 | 0.92 | 1.02 | 1.82×10^-1^ |  | 0.20 | 1.01 | 0.97 | 1.05 | 6.93×10^-1^ |
| rs75295329 | 12q24.11 | 111344621 | T | G |  | 0.13 | 1.12 | 1.06 | 1.19 | 1.27×10^-4^ |  | NA | NA | NA | NA | NA |  | NA | NA | NA | NA | NA |
| rs753955 | 13q12.12 | 24293859 | G | A |  | 0.30 | 1.06 | 1.01 | 1.10 | 1.08×10^-2^ |  | 0.62 | 1.00 | 0.97 | 1.02 | 8.00×10^-1^ |  | 0.49 | 1.04 | 1.01 | 1.07 | 6.12×10^-3^ |
| rs56084662 | 13q13.1 | 32869864 | A | G |  | 0.00 | NA | NA | NA | NA |  | 0.00 | NA | NA | NA | NA |  | NA | NA | NA | NA | NA |
| rs11571833 | 13q13.1 | 32972626 | A | T |  | 0.00 | NA | NA | NA | NA |  | 0.01 | 1.60 | 1.43 | 1.80 | 6.12×10^-16^ |  | NA | NA | NA | NA | NA |
| rs1200399 | 14q13.1 | 35293185 | C | T |  | 0.49 | 1.11 | 1.06 | 1.15 | 4.06×10^-7^ |  | 0.05 | 1.07 | 1.01 | 1.12 | 1.33×10^-2^ |  | 0.41 | 1.11 | 1.07 | 1.15 | 3.05×10^-9^ |
| rs66759488 | 15q21.1 | 47577451 | A | G |  | 0.07 | 0.99 | 0.92 | 1.06 | 7.58×10^-1^ |  | 0.36 | 1.07 | 1.04 | 1.10 | 2.83×10^-8^ |  | 0.68 | 0.96 | 0.93 | 0.99 | 2.30×10^-2^ |
| rs77468143 | 15q21.1 | 49376624 | T | G |  | 0.80 | 1.08 | 1.03 | 1.13 | 1.93×10^-3^ |  | 0.75 | 1.09 | 1.06 | 1.12 | 1.00×10^-9^ |  | 0.76 | 1.09 | 1.05 | 1.12 | 1.24×10^-7^ |
| rs8034191 | 15q25.1 | 78806023 | C | T |  | 0.03 | 1.03 | 0.92 | 1.15 | 6.08×10^-1^ |  | 0.37 | 1.28 | 1.25 | 1.31 | 8.91×10^-96^ |  | 0.32 | 1.25 | 1.20 | 1.29 | 3.29×10^-37^ |
| rs55781567 | 15q25.1 | 78857986 | G | C |  | 0.03 | 1.13 | 1.02 | 1.26 | 2.19×10^-2^ |  | 0.37 | 1.30 | 1.27 | 1.33 | 3.08×10^-103^ |  | 0.32 | 1.27 | 1.23 | 1.31 | 8.44×10^-44^ |
| rs6495306 | 15q25.1 | 78865893 | A | G |  | 0.81 | 1.01 | 0.97 | 1.06 | 5.89×10^-1^ |  | 0.59 | 1.09 | 1.06 | 1.11 | 6.40×10^-13^ |  | 0.67 | 1.06 | 1.03 | 1.09 | 1.54×10^-4^ |
| rs680244 | 15q25.1 | 78871288 | C | T |  | 0.74 | 1.00 | 0.96 | 1.05 | 9.51×10^-1^ |  | 0.58 | 1.09 | 1.06 | 1.12 | 3.51×10^-13^ |  | 0.65 | 1.05 | 1.02 | 1.08 | 7.94×10^-4^ |
| rs16969968 | 15q25.1 | 78882925 | A | G |  | 0.03 | 1.11 | 0.99 | 1.24 | 6.47×10^-2^ |  | 0.36 | 1.29 | 1.26 | 1.32 | 3.32×10^-101^ |  | 0.32 | 1.26 | 1.22 | 1.31 | 1.35×10^-41^ |
| rs1051730 | 15q25.1 | 78894339 | A | G |  | 0.03 | 1.12 | 1.00 | 1.25 | 5.28×10^-2^ |  | 0.36 | 1.29 | 1.26 | 1.32 | 5.52×10^-101^ |  | 0.32 | 1.26 | 1.22 | 1.31 | 8.42×10^-42^ |
| rs938682 | 15q25.1 | 78896547 | A | G |  | 0.55 | 1.02 | 0.98 | 1.06 | 2.96×10^-1^ |  | 0.78 | 1.26 | 1.23 | 1.30 | 3.93×10^-60^ |  | 0.65 | 1.13 | 1.10 | 1.16 | 1.13×10^-16^ |
| rs12914385 | 15q25.1 | 78898723 | T | C |  | 0.28 | 1.03 | 0.98 | 1.07 | 2.49×10^-1^ |  | 0.36 | 1.27 | 1.23 | 1.32 | 1.72×10^-47^ |  | 0.35 | 1.17 | 1.13 | 1.20 | 9.26×10^-29^ |
| rs8042374 | 15q25.1 | 78908032 | A | G |  | 0.30 | 1.01 | 0.97 | 1.06 | 5.44×10^-1^ |  | 0.78 | 1.26 | 1.23 | 1.30 | 2.91×10^-60^ |  | 0.53 | 1.13 | 1.10 | 1.17 | 2.05×10^-16^ |
| rs6495309 | 15q25.1 | 78915245 | C | T |  | 0.55 | 1.02 | 0.98 | 1.06 | 2.73×10^-1^ |  | 0.79 | 1.25 | 1.22 | 1.29 | 6.79×10^-55^ |  | 0.66 | 1.12 | 1.09 | 1.15 | 8.63×10^-15^ |
| rs200595745 | 17q24.2 | 65915289 | A | AAATAATAAT |  | 0.62 | 1.12 | 1.08 | 1.17 | 2.67×10^-8^ |  | 0.23 | 1.06 | 1.02 | 1.11 | 3.33×10^-3^ |  | 0.43 | 1.09 | 1.06 | 1.12 | 2.37×10^-9^ |
| rs7216064 | 17q24.3 | 65898809 | A | G |  | 0.62 | 1.12 | 1.08 | 1.17 | 2.41×10^-8^ |  | 0.20 | 1.04 | 1.01 | 1.07 | 3.40×10^-3^ |  | 0.42 | 1.09 | 1.06 | 1.12 | 1.19×10^-8^ |
| rs56113850 | 19q13.2 | 41353107 | C | T |  | 0.37 | 1.02 | 0.98 | 1.07 | 3.76×10^-1^ |  | 0.56 | 1.13 | 1.10 | 1.16 | 5.02×10^-19^ |  | 0.59 | 0.90 | 0.87 | 0.93 | 2.69×10^-9^ |
| rs6141383 | 20q11.21 | 31889141 | G | A |  | 0.98 | 1.06 | 0.87 | 1.30 | 5.57×10^-1^ |  | 0.00 | NA | NA | NA | NA |  | NA | NA | NA | NA | NA |
| rs4809957 | 20q13.2 | 52771171 | A | G |  | 0.36 | 1.05 | 1.01 | 1.09 | 1.63×10^-2^ |  | 0.79 | 1.03 | 1.00 | 1.06 | 6.93×10^-2^ |  | 0.56 | 1.03 | 1.00 | 1.06 | 6.67×10^-2^ |
| rs41309931 | 20q13.33 | 62326579 | T | G |  | 0.22 | 1.02 | 0.97 | 1.07 | 4.07×10^-1^ |  | 0.12 | 1.08 | 1.04 | 1.12 | 2.23×10^-5^ |  | 0.18 | 1.04 | 1.01 | 1.08 | 1.62×10^-2^ |
| rs17879961 | 22q12.1 | 29121087 | G | A |  | 0.00 | NA | NA | NA | NA |  | 0.99 | 1.66 | 1.42 | 1.94 | 1.54×10^-10^ |  | NA | NA | NA | NA | NA |
| rs36600 | 22q12.2 | 30337586 | T | C |  | 0.10 | 1.01 | 0.94 | 1.08 | 8.18×10^-1^ |  | 0.26 | 1.01 | 0.98 | 1.04 | 4.50×10^-1^ |  | 0.23 | 1.00 | 0.97 | 1.04 | 7.86×10^-1^ |
| rs17728461 | 22q12.2 | 30598552 | G | C |  | 0.19 | 1.06 | 1.01 | 1.11 | 2.40×10^-2^ |  | 0.33 | 1.02 | 1.00 | 1.05 | 7.30×10^-2^ |  | 0.28 | 1.04 | 1.01 | 1.07 | 1.44×10^-2^ |

^†^ EAF: effect allele frequency, derived from the corresponding GWAS datasets.

Table E3. Associations of cross-used ethnic-specific polygenic risk score (PRS) with incident lung cancer in the CKB and the UKB

| Cohorts | Genetic risk ^†^ | No. of cases / Person-years | Model 1^‡^ | |  | Model 2^§^ | |
| --- | --- | --- | --- | --- | --- | --- | --- |
|  |  |  | HR (95% CI) | *P*-value |  | HR (95% CI) | *P*-value |
| CKB | Low | 259/198,206 | Ref |  |  | Ref |  |
|  | Intermediate | 801/594,025 | 1.04 (0.90-1.19) | 0.617 |  | 1.04 (0.90-1.19) | 0.620 |
|  | High | 332/198,218 | 1.32 (1.12-1.56) | 0.001 |  | 1.31 (1.11-1.54) | 0.001 |
|  | *P* value for trend |  | 0.001 | |  | 0.001 | |
| UKB | Low | 327/575,783 | Ref |  |  | Ref |  |
|  | Intermediate | 1197/1,724,811 | 1.22 (1.08-1.38) | 0.001 |  | 1.23 (1.09-1.39) | 0.001 |
|  | High | 501/574,538 | 1.55 (1.35-1.78) | 8.84×10^-10^ |  | 1.57 (1.36-1.80) | 2.98×10^-10^ |
|  | *P* value for trend |  | 4.30×10^-10^ | |  | 1.40×10^-10^ | |

^†^ Genetic risk was categorized into low (the bottom quintile), intermediate (quintiles 2-4) and high (the top quintile) according to distributions of PRSs;

^‡^ Adjusted for age, sex, BMI, highest education level, family cancer history, history of COPD and cancer, lung function and the first 10 principal components of ancestry;

^§^ Additional adjusting for smoking status.

Table E4. Risk of incident lung cancer additionally adjusting for environmental exposures

| Genetic risk ^†^ | Chinese-specific PRS in the CKB | | | | |  | White-specific PRS in the UKB | | | | |
| --- | --- | --- | --- | --- | --- | --- | --- | --- | --- | --- | --- |
|  | No. of cases / Person-years |  | HR (95% CI) ^‡^ |  | *P*-value ^‡^ |  | No. of cases / Person-years |  | HR (95% CI) ^‡^ |  | *P*-value ^‡^ |
| Per SD increase of PRS | 1,392/990,449 |  | 1.19(1.13-1.25) |  | 2.87×10^-10^ |  | 2,025/2,875,132 |  | 1.24(1.19-1.29) |  | 4.97×10^-23^ |
| Low | 221/198,111 |  | Ref |  |  |  | 273/575,454 |  | Ref |  |  |
| Intermediate | 822/594,440 |  | 1.25(1.07-1.45) |  | 0.004 |  | 1,212/1,724,837 |  | 1.43(1.26-1.63) |  | 7.85×10^-8^ |
| High | 349/197,898 |  | 1.62(1.37-1.92) |  | 2.83×10^-8^ |  | 540/574,841 |  | 1.88(1.62-2.17) |  | 2.10×10^-17^ |
| *P*-value for trend |  |  |  |  | 1.46×10^-8^ |  |  |  |  |  | 5.12×10^-18^ |

^†^ Genetic risk were categorized into low (the bottom quintile), intermediate (quintiles 2-4) and high (the top quintile) according to distributions of PRSs；

^‡^ Adjusting for age, sex, smoking status, BMI, highest education level, family history of cancer, personal medical history (previous cancer diagnoses and chronic obstructive pulmonary disease), passive smoking, ambient PM_2.5_ concentration, the forced expiratory volume in 1 second, and the top ten principal components of ancestry.

Table E5. Risk of incident lung cancer according to different risk levels of polygenic risk score (PRS)

| Cohort | Genetic risk | Ethnic-specific PRS | | | |
| --- | --- | --- | --- | --- | --- |
|  |  | No. of cases / Person-years |  | Model ^†^ | |
|  |  |  |  | HR (95% CI) | *P*-value |
| CKB | Quartiles of PRS^‡^ | |  |  |  |
|  | Low | 287/247,519 |  | Ref |  |
|  | Intermediate | 674/495,657 |  | 1.18 (1.03-1.35) | 0.020 |
|  | High | 431/247,273 |  | 1.53 (1.32-1.78) | 2.99×10^-8^ |
|  | *P* value for trend |  |  | 1.53×10^-8^ | |
|  | Tertiles of PRS^§^ |  |  |  |  |
|  | Low | 393/330,058 |  | Ref |  |
|  | Intermediate | 463/330,358 |  | 1.18 (1.03-1.35) | 0.019 |
|  | High | 536/330,033 |  | 1.39 (1.22-1.59) | 7.23×10^-7^ |
|  | *P* value for trend |  |  | 6.42×10^-7^ | |
| UKB | Quartiles of PRS^‡^ |  |  |  |  |
|  | Low | 358/718,903 |  | Ref |  |
|  | Intermediate | 1025/1,437,739 |  | 1.39 (1.24-1.57) | 6.55×10^-8^ |
|  | High | 642/718,490 |  | 1.72 (1.51-1.95) | 2.80×10^-16^ |
|  | *P* value for trend |  |  | 2.24×10^-16^ | |
|  | Tertiles of PRS^§^ |  |  |  |  |
|  | Low | 495/958,532 |  | Ref |  |
|  | Intermediate | 687/958,372 |  | 1.36 (1.22-1.53) | 1.32×10^-7^ |
|  | High | 843/958,228 |  | 1.64 (1.47-1.83) | 2.54×10^-18^ |
|  | *P* value for trend |  |  | 2.62×10^-18^ | |

^†^ Adjusting for age, sex, smoking status, BMI, highest education level, family history of cancer, personal medical history (previous cancer diagnoses and chronic obstructive pulmonary disease), the forced expiratory volume in 1 second, and the top ten principal components of ancestry;

^‡^ Defined by quartiles of composite PRS: low (the bottom quartile), intermediate (quartiles 2-3) and high (the top quartile);

^§^ Defined by tertiles of composite PRS: low (the lowest tertiles), intermediate (the mid tertiles) and high (the highest tertiles).

Table E6. Risk of incident lung cancer after excluding participants with missing covariates

| Cohort | Genetic risk ^†^ | Ethnic-specific PRS | | |
| --- | --- | --- | --- | --- |
|  |  | No. of cases / Person-years | HR (95% CI)^‡^ | *P*-value^‡^ |
| CKB | Low | 206/188,804 | Ref |  |
|  | Intermediate | 760/566,669 | 1.24(1.06-1.44) | 0.007 |
|  | High | 332/188,594 | 1.66(1.39-1.97) | 1.80×10^-8^ |
|  | *P* value for trend | 7.32×10^-9^ | | |
| UKB | Low | 164/380,776 | Ref |  |
|  | Intermediate | 702/1,142,082 | 1.40(1.18-1.66) | 1.01×10^-4^ |
|  | High | 311/380,664 | 1.80(1.49-2.17) | 1.23×10^-9^ |
|  | *P* value for trend | 7.01×10^-10^ | | |

^†^ Genetic risk was categorized into low (the bottom quintile), intermediate (quintiles 2-4) and high (the top quintile) according to distributions of PRSs;

^‡^ Adjusting for age, sex, smoking status, BMI, highest education level, family history of cancer, personal medical history (previous cancer diagnoses and chronic obstructive pulmonary disease), the forced expiratory volume in 1 second, and the top ten principal components of ancestry.

Table E7. Risk of incident lung cancer after excluding incident cases during the first year of follow-up

| Cohort | Genetic risk ^†^ | Ethnic-specific PRS | | |
| --- | --- | --- | --- | --- |
|  |  | No. of cases / Person-years | HR (95% CI)^‡^ | *P*-value^‡^ |
| CKB | Low | 201/198,157 | Ref |  |
|  | Intermediate | 783/594,339 | 1.31(1.12-1.53) | 0.001 |
|  | High | 331/197,911 | 1.69(1.41-2.02) | 6.56×10^-9^ |
|  | *P* value for trend | 3.67×10^-9^ | | |
| UKB | Low | 248/575,330 | Ref |  |
|  | Intermediate | 1101/1,724,734 | 1.44(1.25-1.65) | 2.44×10^-7^ |
|  | High | 480/574,958 | 1.84(1.57-2.14) | 8.57×10^-15^ |
|  | *P* value for trend | 4.24×10^-15^ | | |

^†^ Genetic risk were categorized into low (the bottom quintile), intermediate (quintiles 2-4) and high (the top quintile) according to distributions of PRSs;

^‡^ Adjusting for age, sex, smoking status, BMI, highest education level, family history of cancer, personal medical history (previous cancer diagnoses and chronic obstructive pulmonary disease), the forced expiratory volume in 1 second, and the top ten principal components of ancestry.

Table E8. Risk of incident lung cancer according to levels of pack-years of smoking in the CKB and the UKB

| Cohort | Smoking status ^†^ | No. of cases / Person-years | Model 1 ^‡^ | |  | Model 2 ^§^ | |
| --- | --- | --- | --- | --- | --- | --- | --- |
|  |  |  | HR (95% CI) | *P* value |  | HR (95% CI) | *P* value |
| CKB | Nonsmoker | 583/662,908 | Ref |  |  | Ref |  |
|  | Light Smoker | 332/208,388 | 1.69 (1.42-2.02) | 3.44×10^-9^ |  | 1.70 (1.43-2.02) | 2.94×10^-9^ |
|  | Heavy Smoker | 477/119,153 | 2.87 (2.40-3.44) | 7.36×10^-31^ |  | 2.88 (2.41-3.45) | 5.57×10^-31^ |
|  | *P* value for trend |  | 3.85×10^-32^ | |  | 4.59×10^-32^ | |
| UKB | Nonsmoker | 289/1,583,534 | Ref |  |  | Ref |  |
|  | Light Smoker | 751/1,049,471 | 3.58 (3.12-4.10) | 4.39×10^-75^ |  | 3.59 (3.14-4.12) | 1.38×10^-75^ |
|  | Heavy Smoker | 985/242,126 | 15.79 (13.77-18.10) | < 1×10^-125^ |  | 15.65 (13.65-17.94) | < 1×10^-125^ |
|  | *P* value for trend |  | < 1×10^-125^ | |  | < 1×10^-125^ | |

^†^ Participants were defined as nonsmokers (less than 100 cigarettes in lifetime), light smokers (pack years of smoking, PY<30), and heavy smokers (PY≥30);

^‡^ Adjusting for age, sex, BMI, highest education level, family history of cancer, personal medical history (previous cancer diagnoses and chronic obstructive pulmonary disease), the forced expiratory volume in 1 second, and the top ten principal components of ancestry.

^§^ Adjusting for age, sex, BMI, highest education level, family history of cancer, personal medical history (previous cancer diagnoses and chronic obstructive pulmonary disease), the forced expiratory volume in 1 second, and the top ten principal components of ancestry, and ethnic-specific PRSs.

Table E9. Risk of incident lung cancer according to smoking status in the CKB and the UKB

| Cohort | Smoking status | No. of cases / Person-years | Model 1^†^ | |  | Model 2^‡^ | |
| --- | --- | --- | --- | --- | --- | --- | --- |
|  |  |  | HR (95% CI) | *P* value |  | HR (95% CI) | *P* value |
| CKB | Never smoker | 583/662,908 | Ref |  |  | Ref |  |
|  | Former smoker | 70/30,484 | 1.59 (1.21-2.09) | 0.001 |  | 1.59 (1.21-2.09) | 0.001 |
|  | Current smoker | 739/297,057 | 2.18 (1.86-2.55) | 5.10×10^-22^ |  | 2.18 (1.86-2.56) | 3.96×10^-22^ |
|  | *P* value for trend | | 3.06×10^-22^ | |  | 2.41×10-22 | |
| UKB | Never smoker | 271/1,577,029 | Ref |  |  | Ref |  |
|  | Former smoker | 932/1,009,064 | 4.17 (3.64-4.78) | 3.19×10^-93^ |  | 4.18 (3.64-4.79) | 1.82×10^-93^ |
|  | Current smoker | 822/289,039 | 15.72 (13.67-18.09) | < 1×10^-125^ |  | 15.78 (13.72-18.16) | < 1×10^-125^ |
|  | *P* value for trend | | < 1×10^-125^ | |  | < 1×10^-125^ | |

^†^ Model1: Adjusting for age, sex, BMI, highest education level, family history of cancer, personal medical history (previous cancer diagnoses and chronic obstructive pulmonary disease), the forced expiratory volume in 1 second, and the top ten principal components of ancestry;

^‡^ Model2: Adjusting for age, sex, BMI, highest education level, family history of cancer, personal medical history (previous cancer diagnoses and chronic obstructive pulmonary disease), the forced expiratory volume in 1 second, and the top ten principal components of ancestry, and ethnic-specific PRSs.

Table E10. Associations between polygenic risk scores (PRSs) and pack-years of smoking

| Cohort | PRS | β (95%CI) ^†^ | *P*-value ^†^ |
| --- | --- | --- | --- |
| CKB | Chinese-specific PRS | -0.022 (-0.113-0.070) | 0.640 |
|  | Trans-ancestry PRS | 0.136 (-0.156-0.428) | 0.360 |
| UKB | White-specific PRS | 0.515 (0.374-0.655) | 7.04×10^-13^ |
|  | Trans-ancestry PRS | 0.336 (0.200-0.473) | 1.27×10^-6^ |

^†^ Adjusted for age, sex, BMI, education level, family cancer history, and history of COPD and cancer.

Table E11. Mediation analysis of pack-years of smoking on the associations between polygenic risk scores (PRSs) and lung cancer

| Cohort | PRS | Total association  β (95%CI) ^†^ | Direct association  β (95%CI) ^†^ | Indirect association  β (95%CI) ^†^ | Mediation proportion  % (95%CI) ^‡^ |
| --- | --- | --- | --- | --- | --- |
| CKB | Chinese-specific PRS | 0.476 (0.325-0.631) | 0.477 (0.329-0.625) | -0.001 (-0.004-0.002) | -0.15 (-0.84-0.47) |
|  | Trans-ancestry PRS | 0.552 (0.384-0.718) | 0.551 (0.381-0.720) | 0.002 (-0.002-0.005) | 0.28 (-0.33-0.98) |
| UKB | White-specific PRS | 0.676 (0.543-0.810) | 0.662 (0.533-0.792) | 0.014 (0.010-0.018) | 2.06 (1.41-2.90) |
|  | Trans-ancestry PRS | 0.640 (0.504-0.768) | 0.630 (0.504-0.757) | 0.009 (0.005-0.013) | 1.43 (0.83-2.22) |

^†^Adjusting for age, sex, BMI, highest education level, family history of cancer, personal medical history (previous cancer diagnoses and chronic obstructive pulmonary disease), the forced expiratory volume in 1 second, and the top ten principal components of ancestry;

^‡^ Generated by drawing 5000 bootstrap samples from the estimation dataset.

Table E12. Interactions between Trans-ancestry polygenic risk scores (PRSs) and pack-years of smoking on the risk of incident lung cancer in the CKB and the UKB

| PRS ^§^ | Additive interaction ^†^ | | | | |
| --- | --- | --- | --- | --- | --- |
|  | Light Smoker (pack-year <30) ^††^ | |  | Heavy Smoker (pack-year ≥30) ^††^ | |
|  | RERI^‡^ (95%CI) | AP^‡^ (95%CI) |  | RERI^‡^ (95%CI) | AP^‡^ (95%CI) |
| CKB |  |  |  |  |  |
| Intermediate | -0.04(-0.71-0.50) | -0.02(-0.36-0.26) |  | -0.06(-0.97-0.74) | -0.02(-0.29-0.21) |
| High | 0.67(-0.16-1.53) | 0.22(-0.06-0.43) |  | 0.57(-0.57-1.74) | 0.13(-0.14-0.34) |
| UKB |  |  |  |  |  |
| Intermediate | 0.27(-0.96-1.34) | 0.05(-0.15-0.23) |  | 7.46(3.51-12.88) | 0.28(0.14-0.41) |
| High | 1.64(0.12-3.24) | 0.22(0.02-0.38) |  | 14.86(8.94-23.81) | 0.43(0.31-0.54) |

^†^ Adjusting for age, sex, BMI, highest education level, family history of cancer, personal medical history (previous cancer diagnoses and chronic obstructive pulmonary disease), the forced expiratory volume in 1 second, and the top ten principal components of ancestry;

^‡^ To estimate RERI and AP, the nonsmoker category and the lowest genetic risk (low PRS) groups were the reference categories;

^§^ Genetic risk was categorized into low (the bottom quintile), intermediate (quintiles 2-4) and high (the top quintile) according to distributions of PRSs;

^††^ Participants were defined as nonsmokers (less than 100 cigarettes in lifetime), light smokers (pack years of smoking, PY<30), and heavy smokers (PY≥30).

Table E13. Interactions between genetic risk and pack-years of smoking on the risk of incident lung cancer additionally adjusting for environmental exposures

| PRS ^§^ | Additive interaction ^†^ | | | | |
| --- | --- | --- | --- | --- | --- |
|  | Light Smoker (pack-year <30) ^††^ | |  | Heavy Smoker (pack-year≥30) ^††^ | |
|  | RERI ^‡^ (95%CI) | AP ^‡^ (95%CI) |  | RERI ^‡^ (95%CI) | AP ^‡^ (95%CI) |
| CKB |  |  |  |  |  |
| Chinese-specific PRS |  |  |  |  |  |
| Intermediate | 0.14 (-0.55 to 0.74) | 0.06 (-0.24 to 0.30) |  | -0.04 (-0.99 to 0.86) | -0.01 (-0.26 to 0.21) |
| High | 0.24 (-0.70 to 1.14) | 0.08 (-0.27 to 0.34) |  | 0.50 (-0.75 to 1.82) | 0.10 (-0.16 to 0.32) |
| UKB |  |  |  |  |  |
| White-specific PRS |  |  |  |  |  |
| Intermediate | 0.79 (0.04 to 1.49) | 0.19 (0.01 to 0.36) |  | 5.25 (3.04 to 8.40) | 0.35 (0.21 to 0.47) |
| High | 2.02 (0.96 to 3.27) | 0.34 (0.17 to 0.49) |  | 8.09 (4.99 to 12.68) | 0.44 (0.30 to 0.55) |

^†^ Adjusting for age, sex, BMI, highest education level, family history of cancer, personal medical history (previous cancer diagnoses and chronic obstructive pulmonary disease), passive smoking, ambient PM_2.5_ concentration, the forced expiratory volume in 1 second, and the top ten principal components of ancestry;

^‡^ To estimate RERI and AP, the nonsmoker category and the lowest genetic risk (low PRS) groups were the reference categories;

^§^ Genetic risk were categorized into low (the bottom quintile), intermediate (quintiles 2-4) and high (the top quintile) according to distributions of PRSs;

^††^ Participants were defined as nonsmokers (less than 100 cigarettes in lifetime), light smokers (pack years of smoking <30), and heavy smokers (pack years of smoking≥30).

Table E14. Interactions between genetic risk and pack-years of smoking on the risk of incident lung cancer according to different risk levels of polygenic risk scores (PRSs) in CKB and UKB cohorts

| Cohort | Genetic risk | Additive interaction ^†^ | | | | |
| --- | --- | --- | --- | --- | --- | --- |
|  |  | Light Smoker (pack-year <30) ^ll^ | |  | Heavy Smoker (pack-year ≥30) ^ll^ | |
|  |  | RERI ^‡^ (95%CI) | AP ^‡^ (95%CI) |  | RERI ^‡^ (95%CI) | AP ^‡^ (95%CI) |
| CKB | Chinese-specific PRS |  |  |  |  |  |
|  | Quartiles of PRS^§^ |  |  |  |  |  |
|  | Intermediate | 0.15 (-0.43-0.67) | 0.07 (-0.22-0.31) |  | 0.06 (-0.79-0.84) | 0.02 (-0.23-0.23) |
|  | High | 0.51 (-0.22-1.28) | 0.18 (-0.09-0.40) |  | 0.57 (-0.48-1.61) | 0.13 (-0.12-0.32) |
|  | Tertiles of PRS^††^ |  |  |  |  |  |
|  | Intermediate | 0.10 (-0.50-0.66) | 0.05 (-0.25-0.28) |  | -0.22 (-1.09-0.60) | -0.06 (-0.34-0.16) |
|  | High | 0.38 (-0.27-0.99) | 0.15 (-0.12-0.34) |  | 0.14 (-0.75-1.02) | 0.03 (-0.20-0.23) |
|  | Trans-ancestry PRS |  |  |  |  |  |
|  | Quartiles of PRS^§^ |  |  |  |  |  |
|  | Intermediate | -0.06 (-0.64-0.47) | -0.03 (-0.35-0.24) |  | -0.13 (-0.98-0.60) | -0.04 (-0.30-0.18) |
|  | High | 0.60 (-0.13-1.33) | 0.21 (-0.05-0.41) |  | 0.60 (-0.41-1.64) | 0.14 (-0.11-0.33) |
|  | Tertiles of PRS^††^ |  |  |  |  |  |
|  | Intermediate | -0.16 (-0.76-0.39) | -0.09 (-0.46-0.19) |  | -0.02 (-0.83-0.74) | -0.01 (-0.28-0.21) |
|  | High | 0.22 (-0.42-0.84) | 0.09 (-0.17-0.29) |  | 0.75 (-0.11-1.67) | 0.17 (-0.03-0.33) |
| UKB | White-specific PRS |  |  |  |  |  |
|  | Quartiles of PRS^§^ |  |  |  |  |  |
|  | Intermediate | 1.11 (0.33-1.89) | 0.23 (0.07-0.38) |  | 5.99 (2.81-9.61) | 0.28 (0.14-0.40) |
|  | High | 1.91 (0.88-3.02) | 0.31 (0.15-0.45) |  | 9.19 (5.29-14.24) | 0.36 (0.23-0.47) |
|  | Tertiles of PRS^††^ |  |  |  |  |  |
|  | Intermediate | 1.37 (0.65-2.18) | 0.30 (0.15-0.44) |  | 5.06 (2.20-8.33) | 0.26 (0.12-0.38) |
|  | High | 1.78 (0.99-2.69) | 0.33 (0.19-0.45) |  | 8.05 (5.03-11.95) | 0.35 (0.24-0.45) |
|  | Trans-ancestry PRS |  |  |  |  |  |
|  | Quartiles of PRS^§^ |  |  |  |  |  |
|  | Intermediate | 0.22 (-0.92-1.19) | 0.04 (-0.15-0.20) |  | 6.90 (3.14-11.78) | 0.26 (0.13-0.38) |
|  | High | 1.45 (0.14-2.83) | 0.20 (0.02-0.35) |  | 10.33 (5.62-16.78) | 0.34 (0.21-0.45) |
|  | Tertiles of PRS^††^ |  |  |  |  |  |
|  | Intermediate | 0.02 (-1.09-1.00) | 0.00 (-0.20-0.18) |  | 7.30 (3.65-11.66) | 0.28 (0.15-0.39) |
|  | High | 1.00 (-0.09-2.10) | 0.15 (-0.01-0.29) |  | 9.01 (5.13-14.13) | 0.32 (0.20-0.42) |

^†^ Adjusting for age, sex, BMI, highest education level, family history of cancer, personal medical history (previous cancer diagnoses and chronic obstructive pulmonary disease), the forced expiratory volume in 1 second, and the top ten principal components of ancestry;

^‡^ To estimate RERI and AP, the Nonsmoker category and the lowest genetic risk (low PRS) groups were the reference categories.

^§^ Defined by quartiles of composite PRS: low (the bottom quartile), intermediate (quartiles 2-3) and high (the top quartile);

^††^ Defined by tertiles of composite PRS: low (the lowest tertiles), intermediate (the mid tertiles) and high (the highest tertiles);

^ll^ Participants were defined as nonsmokers (less than 100 cigarettes in lifetime), light smokers (pack years of smoking, PY<30), and heavy smokers (PY≥30)

Table E15. Interactions between genetic risk and pack-years of smoking on the risk of incident lung cancer after excluding incident cases during the first year of follow-up

| PRS ^§^ | Additive interaction ^†^ | | | | |
| --- | --- | --- | --- | --- | --- |
|  | Light Smoker (pack-year <30) ^††^ | |  | Heavy Smoker (pack-year ≥30) ^††^ | |
|  | RERI^‡^ (95%CI) | AP^‡^ (95%CI) |  | RERI^‡^ (95%CI) | AP^‡^ (95%CI) |
| CKB |  |  |  |  |  |
| Chinese-specific PRS |  |  |  |  |  |
| Intermediate | 0.18 (-0.57-0.84) | 0.07 (-0.22-0.31) |  | -0.11 (-1.24-0.93) | -0.03 (-0.30-0.21) |
| High | 0.17 (-0.88-1.18) | 0.05 (-0.32-0.32) |  | 0.54 (-0.84-1.97) | 0.10 (-0.18-0.32) |
| Trans-ancestry PRS |  |  |  |  |  |
| Intermediate | -0.04 (-0.74-0.56) | -0.02 (-0.36-0.27) |  | -0.16 (-1.22-0.72) | -0.05 (-0.34-0.20) |
| High | 0.69 (-0.24-1.67) | 0.21 (-0.09-0.43) |  | 0.47 (-0.79-1.74) | 0.10 (-0.18-0.32) |
| UKB |  |  |  |  |  |
| White-specific PRS |  |  |  |  |  |
| Intermediate | 1.01 (0.14-1.86) | 0.22 (0.03-0.40) |  | 7.32 (3.98-11.80) | 0.34 (0.20-0.47) |
| High | 2.09 (0.90-3.53) | 0.34 (0.15-0.49) |  | 10.96 (6.47-17.64) | 0.43 (0.29-0.55) |
| Trans-ancestry PRS |  |  |  |  |  |
| Intermediate | 0.41 (-0.91-1.62) | 0.07 (-0.13-0.25) |  | 8.46 (4.08-15.12) | 0.29 (0.16-0.42) |
| High | 1.86 (0.25-3.83) | 0.23 (0.03-0.40) |  | 15.17 (8.77-26.07) | 0.42 (0.29-0.54) |

^†^ Adjusting for age, sex, BMI, highest education level, family history of cancer, personal medical history (previous cancer diagnoses and chronic obstructive pulmonary disease), the forced expiratory volume in 1 second, and the top ten principal components of ancestry;

^‡^ To estimate RERI and AP, the Nonsmoker category and the lowest genetic risk (low PRS) groups were the reference categories.

^§^ Genetic risk was categorized into low (the bottom quintile), intermediate (quintiles 2-4) and high (the top quintile) according to distributions of PRSs;

^††^ Participants were defined as nonsmokers (less than 100 cigarettes in lifetime), light smokers (pack years of smoking, PY<30), and heavy smokers (PY≥30).

Table E16. Interactions between genetic risk and pack-years of smoking on the risk of incident lung cancer after excluding participants with missing covariates

| PRS ^§^ | Additive interaction ^†^ | | | | |
| --- | --- | --- | --- | --- | --- |
|  | Light Smoker (pack-year<30) ^††^ | |  | Heavy Smoker (pack-year ≥30) ^††^ | |
|  | RERI^‡^ (95%CI) | AP^‡^ (95%CI) |  | RERI^‡^ (95%CI) | AP^‡^ (95%CI) |
| CKB |  |  |  |  |  |
| Chinese-specific PRS |  |  |  |  |  |
| Intermediate | 0.21 (-0.55-0.83) | 0.09 (-0.23-0.34) |  | -0.15 (-1.27-0.78) | -0.04 (-0.33-0.20) |
| High | 0.20 (-0.80-1.15) | 0.07 (-0.30-0.33) |  | 0.51 (-0.83-1.88) | 0.10 (-0.18-0.32) |
| Trans-ancestry PRS |  |  |  |  |  |
| Intermediate | -0.06 (-0.73-0.51) | -0.03 (-0.38-0.26) |  | 0.05 (-0.91-0.87) | 0.01 (-0.27-0.25) |
| High | 0.65 (-0.24-1.57) | 0.20 (-0.09-0.42) |  | 0.70 (-0.55-1.97) | 0.15 (-0.13-0.36) |
| UKB |  |  |  |  |  |
| White-specific PRS |  |  |  |  |  |
| Intermediate | 0.63 (-0.43-1.52) | 0.16 (-0.10-0.39) |  | 6.47 (3.25-10.84) | 0.40 (0.22-0.54) |
| High | 2.15 (0.84-3.81) | 0.37 (0.16-0.55) |  | 8.43 (4.36-14.29) | 0.45 (0.27-0.60) |
| Trans-ancestry PRS |  |  |  |  |  |
| Intermediate | -0.06 (-1.87-1.28) | -0.01 (-0.28-0.22) |  | 5.74 (1.25-11.98) | 0.25 (0.06-0.42) |
| High | 1.75 (-0.23-4.09) | 0.22 (-0.03-0.42) |  | 11.55 (5.33-22.30) | 0.40 (0.22-0.55) |

^†^ Adjusting for age, sex, BMI, highest education level, family history of cancer, personal medical history (previous cancer diagnoses and chronic obstructive pulmonary disease), the forced expiratory volume in 1 second, and the top ten principal components of ancestry;

^‡^ To estimate RERI and AP, the Nonsmoker category and the lowest genetic risk (low PRS) groups were the reference categories;

^§^ Genetic risk was categorized into low (the bottom quintile), intermediate (quintiles 2-4) and high (the top quintile) according to distributions of PRSs;

^††^ Participants were defined as nonsmokers (less than 100 cigarettes in lifetime), light smokers (pack years of smoking, PY<30), and heavy smokers (PY≥30).

Table E17. Risk of incident cancer according to pack-years of smoking within each genetic risk (quartiles) level in the CKB and the UKB ^†^

| Cohort | PRS | Smoking status | Low genetic risk | | |  | Intermediate genetic risk | | |  | High genetic risk | | |
| --- | --- | --- | --- | --- | --- | --- | --- | --- | --- | --- | --- | --- | --- |
|  |  |  | Heavy Smoker | Light Smoker | Nonsmoker |  | Heavy Smoker | Light Smoker | Nonsmoker |  | Heavy Smoker | Light Smoker | Nonsmoker |
| CKB | Chinese-specific PRS | No. of cases/ Person years | 105/29394 | 69/54148 | 113/163977 |  | 228/59498 | 161/104183 | 285/331977 |  | 144/30261 | 102/50058 | 185/166954 |
|  |  | Hazards ratio (95%CI) ^‡^ | Ref. | 0.50 (0.37-0.69) | 0.24 (0.16-0.36) |  | Ref. | 0.59 (0.48-0.72) | 0.34 (0.26-0.44) |  | Ref. | 0.67 (0.51-0.87) | 0.47 (0.34-0.64) |
|  |  | *P* value |  | 1.84×10^-5^ | 2.60×10^-12^ |  |  | 5.07×10^-7^ | 2.03×10^-16^ |  |  | 2.37×10^-3^ | 1.68×10^-6^ |
|  |  | *P* value for trend | 9.22×10^-13^ | | |  | 5.57×10^-17^ | | |  | 9.56×10^-7^ | | |
|  |  | Absolute risk (%)-5 years (95% CI) ^§^ | 9.22 (6.84-11.60) | 4.58 (3.28-5.89) | 2.52 (1.90-3.13) |  | 9.93 (8.21-11.64) | 5.45 (4.43-6.47) | 3.01 (2.53-3.48) |  | 11.63 (9.09-14.18) | 7.03 (5.37-8.70) | 3.71 (2.97-4.45) |
|  |  | Absolute risk reduction (%)-5 years (95% CI) ^§^ | Ref. | 4.64 (2.03-6.82) | 6.70 (4.04-8.82) |  | Ref. | 4.48 (2.55-6.09) | 6.92 (5.19-8.46) |  | Ref. | 4.60 (1.95-7.08) | 7.93 (5.30-10.13) |
|  | Trans-ancestry PRS | No. of cases/ Person years | 107/29342 | 72/53485 | 117/164774 |  | 223/59289 | 151/104023 | 289/332389 |  | 147/30522 | 109/50881 | 177/165744 |
|  |  | Hazards ratio (95%CI) ^‡^ | Ref. | 0.51 (0.37-0.70) | 0.25 (0.17-0.38) |  | Ref. | 0.57 (0.46-0.70) | 0.36 (0.28-0.47) |  | Ref. | 0.67 (0.52-0.87) | 0.41 (0.30-5.59) |
|  |  | *P* value |  | 2.13×10^-5^ | 1.19×10^-11^ |  |  | 2.16×10^-7^ | 7.72×10^-15^ |  |  | 2.29×10^-3^ | 2.91×10^-8^ |
|  |  | *P* value for trend | 4.04×10^-12^ | | |  | 1.44×10^-15^ | | |  | 2.03×10^-8^ | | |
|  |  | Absolute risk (%)-5 years (95% CI) ^§^ | 9.62 (7.18-12.05) | 4.92 (3.55-6.29) | 2.64 (2.01-3.27) |  | 9.49 (7.82-11.16) | 5.04 (4.07-6.01) | 2.98 (2.51-3.46) |  | 12.03 (9.43-14.64) | 7.40 (5.70-9.11) | 3.63 (2.90-4.36) |
|  |  | Absolute risk reduction (%)-5 years (95% CI) ^§^ | Ref. | 4.69 (2.32-7.18) | 6.98 (4.54-9.20) |  | Ref. | 4.45 (2.75-6.09) | 6.51 (4.94-7.98) |  | Ref. | 4.63 (2.09-7.18) | 8.40 (5.84-10.71) |
| UKB | White-specific PRS | No. of cases/ Person years | 167/56545 | 137/265895 | 54/396464 |  | 506/120925 | 380/526401 | 139/790413 |  | 312/64657 | 234/257176 | 96/396657 |
|  |  | Hazards ratio (95%CI) ^‡^ | Ref. | 0.22 (0.17-0.28) | 0.06 (0.05-0.09) |  | Ref. | 0.23 (0.20-0.26) | 0.06 (0.05-0.07) |  | Ref. | 0.23 (0.20-0.28) | 0.07 (0.05-0.09) |
|  |  | *P* value |  | 4.07×10^-36^ | 2.15×10^-63^ |  |  | 3.83×10^-97^ | 5.34×10^-172^ |  |  | 7.09×10^-59^ | 4.19×10^-109^ |
|  |  | *P* value for trend | 3.75×10^-69^ | | |  | 1.06×10^-195^ | | |  | 1.53×10^-120^ | | |
|  |  | Absolute risk (%)-5 years (95% CI) ^§^ | 8.37 (6.63-10.12) | 1.72 (1.36-2.08) | 0.49 (0.35-0.64) |  | 11.35 (9.95-12.74) | 2.33 (2.03-2.62) | 0.61 (0.50-0.72) |  | 14.87 (12.65-17.08) | 3.22 (2.72-3.73) | 0.91 (0.71-1.11) |
|  |  | Absolute risk reduction (%)-5 years (95% CI) ^§^ | Ref. | 6.65 (4.92-8.15) | 7.88 (6.01-9.52) |  | Ref. | 9.02 (7.61-10.41) | 10.74 (9.28-12.22) |  | Ref. | 11.64 (9.40-13.69) | 13.96 (11.64-16.13) |
|  | Trans-ancestry PRS | No. of cases/ Person years | 173/57501 | 156/267265 | 44/394203 |  | 509/120264 | 366/525899 | 152/791817 |  | 303/64361 | 229/256308 | 93/397514 |
|  |  | Hazards ratio (95%CI) ^‡^ | Ref. | 0.27 (0.21-0.33) | 0.06 (0.04-0.08) |  | Ref. | 0.21 (0.18-0.24) | 0.06 (0.05-0.08) |  | Ref. | 0.24 (0.20-0.28) | 0.07 (0.05-0.09) |
|  |  | *P* value |  | 1.79×10^-30^ | 2.35×10^-59^ |  |  | 5.17×10^-106^ | 1.10×10^-180^ |  |  | 2.43×10^-56^ | 4.72×10^-105^ |
|  |  | *P* value for trend | 8.30×10-^70^ | | |  | 3.68×10^-200^ | | |  | 4.30×10^-116^ | | |
|  |  | Absolute risk (%)-5 years (95% CI) ^§^ | 7.60 (5.99-9.21) | 1.76 (1.40-2.13) | 0.37 (0.25-0.48) |  | 12.38 (10.90-13.87) | 2.40 (2.10-2.70) | 0.71 (0.59-0.83) |  | 13.79 (11.67-15.90) | 3.00 (2.52-3.48) | 0.84 (0.66-1.03) |
|  |  | Absolute risk reduction (%)-5 years (95% CI) ^§^ | Ref. | 5.84 (4.17-7.29) | 7.23 (5.53-8.71) |  | Ref. | 9.98 (8.51-11.38) | 11.67 (10.18-13.11) |  | Ref. | 10.78 (8.69-12.57) | 12.94 (10.87-14.96) |

^†^ Genetic risk was defined by quartiles of PRS: low (the bottom quartile), intermediate (quartiles 2-4) and high (the top quartile);

^‡^ The HRs were estimated using Cox proportional hazards regression with adjustment for age, sex, BMI, highest education level, family history of cancer, personal medical history (previous cancer diagnoses and chronic obstructive pulmonary disease), the forced expiratory volume in 1 second, and the top ten principal components of ancestry;

^§^ The 5-year absolute risk reduction and 95% CI were generated by drawing 1000 bootstrap samples from the estimation dataset.

Table E18. Risk of incident cancer according to pack-years of smoking within each genetic risk (tertiles) level in the CKB and the UKB ^†^

| Cohort | PRS | Smoking status | Low genetic risk | | |  | Intermediate genetic risk | | |  | High genetic risk | | |
| --- | --- | --- | --- | --- | --- | --- | --- | --- | --- | --- | --- | --- | --- |
|  |  |  | Heavy Smoker | Light Smoker | Nonsmoker |  | Heavy Smoker | Light Smoker | Nonsmoker |  | Heavy Smoker | Light Smoker | Nonsmoker |
| CKB | Chinese-specific PRS | No. of cases/ Person years | 149/39654 | 92/71168 | 152/219235 |  | 153/39412 | 111/69830 | 199/221116 |  | 175/40086 | 129/67391 | 232/222556 |
|  |  | Hazards ratio (95%CI) ^‡^ | Ref. | 0.48 (0.37-0.63) | 0.24 (0.17-3.30) |  | Ref. | 0.60 (0.47-0.78) | 0.36 (0.26-0.49) |  | Ref. | 0.67 (0.53-0.84) | 0.45 (0.34-0.60) |
|  |  | *P* value |  | 1.05×10^-7^ | 5.72×10^-17^ |  |  | 8.94×10^-5^ | 1.59×10^-10^ |  |  | 7.22×10^-4^ | 2.93×10^-8^ |
|  |  | *P* value for trend | 8.14×10^-18^ | | |  | 7.62×10^-11^ | | |  | 1.60×10^-8^ | | |
|  |  | Absolute risk (%)-5 years (95% CI) ^§^ | 9.94 (7.78-12.10) | 4.76 (3.59-5.92) | 2.57 (2.03-3.11) |  | 9.67 (7.62-11.72) | 5.41 (4.18-6.64) | 3.06 (2.47-3.64) |  | 10.93 (8.78-13.08) | 6.65 (5.25-8.05) | 3.53 (2.91-4.16) |
|  |  | Absolute risk reduction (%)-5 years (95% CI) ^§^ | Ref. | 5.18 (2.93-7.10) | 7.37 (5.24-9.24) |  | Ref. | 4.26 (2.13-6.24) | 6.61 (4.68-8.42) |  | Ref. | 4.28 (2.09-6.18) | 7.40 (5.32-9.21) |
|  | Trans-ancestry PRS | No. of cases/ Person years | 137/39117 | 102/71742 | 157/219377 |  | 143/39324 | 97/68940 | 183/222008 |  | 197/40712 | 133/67706 | 243/221523 |
|  |  | Hazards ratio (95%CI) ^‡^ | Ref. | 0.57 (0.44-0.75) | 0.27 (0.19-0.38) |  | Ref. | 0.56 (0.43-0.73) | 0.33 (0.24-0.45) |  | Ref. | 0.61 (0.49-0.77) | 0.44 (0.33-0.57) |
|  |  | *P* value |  | 4.19×10^-5^ | 3.55×10^-14^ |  |  | 1.95×10^-5^ | 1.40×10^-11^ |  |  | 2.39×10^-5^ | 2.71×10^-9^ |
|  |  | *P* value for trend | 2.55×10^-14^ | | |  | 4.91×10^-12^ | | |  | 7.00×10^-10^ | | |
|  |  | Absolute risk (%)-5 years (95% CI) ^§^ | 8.65 (6.70-10.60) | 4.95 (3.77-6.13) | 2.52 (1.99-3.04) |  | 9.74 (7.63-11.85) | 5.14 (3.92-6.36) | 2.99 (2.41-3.58) |  | 12.02 (9.75-14.28) | 6.70 (5.32-8.09) | 3.66 (3.03-4.30) |
|  |  | Absolute risk reduction (%)-5 years (95% CI) ^§^ | Ref. | 3.70 (1.74-5.43) | 6.13 (4.11-7.91) |  | Ref. | 4.60 (2.24-6.48) | 6.75 (4.65-8.55) |  | Ref. | 5.31 (2.99-7.45) | 8.35 (6.17-10.29) |
| UKB | White-specific PRS | No. of cases/ Person years | 235/76415 | 181/354187 | 79/527930 |  | 334/80250 | 265/351599 | 88/526524 |  | 416/85462 | 305/343686 | 122/529080 |
|  |  | Hazards ratio (95%CI) ^‡^ | Ref. | 0.22 (0.18-0.26) | 0.07 (0.05-0.09) |  | Ref. | 0.24 (0.20-0.28) | 0.06 (0.05-0.07) |  | Ref. | 0.23 (0.20-0.27) | 0.07 (0.05-0.08) |
|  |  | *P* value |  | 3.98×10^-50^ | 2.58×10^-85^ |  |  | 7.90×10^-64^ | 3.17×10^-116^ |  |  | 2.53×10^-78^ | 1.19×10^-142^ |
|  |  | *P* value for trend | 1.21×10^-91^ | | |  | 1.54×10^-134^ | | |  | 2.97×10^-159^ | | |
|  |  | Absolute risk (%)-5 years (95% CI) ^§^ | 8.37 (6.87-9.87) | 1.65 (1.35-1.96) | 0.52 (0.40-0.65) |  | 11.65 (9.92-13.38) | 2.48 (2.11-2.85) | 0.59 (0.46-0.72) |  | 14.43 (12.53-16.32) | 3.06 (2.64-3.48) | 0.85 (0.69-1.01) |
|  |  | Absolute risk reduction (%)-5 years (95% CI) ^§^ | Ref. | 6.72 (5.20-8.07) | 7.85 (6.31-9.31) |  | Ref. | 9.17 (7.50-10.89) | 11.06 (9.33-12.82) |  | Ref. | 11.37 (9.53-13.25) | 13.58 (11.73-15.58) |
|  | Trans-ancestry PRS | No. of cases/ Person years | 237/76884 | 216/356732 | 62/524964 |  | 353/80777 | 238/350111 | 100/527691 |  | 395/84465 | 297/342628 | 127/530879 |
|  |  | Hazards ratio (95%CI) ^‡^ | Ref. | 0.27 (0.22-0.32) | 0.06 (0.04-0.08) |  | Ref. | 0.20 (0.17-0.23) | 0.06 (0.05-0.08) |  | Ref. | 0.23 (0.20-0.27) | 0.07 (0.06-0.09) |
|  |  | *P* value |  | 2.52×10^-41^ | 6.10×10^-82^ |  |  | 3.88×10^-77^ | 5.06×10^-126^ |  |  | 1.81×10^-74^ | 4.79×10^-137^ |
|  |  | *P* value for trend | 6.65×10^-96^ | | |  | 1.24×10^-140^ | | |  | 9.36×10^-150^ | | |
|  |  | Absolute risk (%)-5 years (95% CI) ^§^ | 7.89 (6.47-9.31) | 1.86 (1.53-2.18) | 0.39 (0.29-0.50) |  | 12.89 (11.03-14.75) | 2.35 (1.99-2.71) | 0.70 (0.55-0.85) |  | 13.79 (11.94-15.65) | 2.96 (2.55-3.38) | 0.88 (0.71-1.05) |
|  |  | Absolute risk reduction (%)-5 years (95% CI) ^§^ | Ref. | 6.04 (4.74-7.31) | 7.50 (6.12-8.89) |  | Ref. | 10.54 (8.51-12.11) | 12.19 (10.11-13.81) |  | Ref. | 10.83 (8.97-12.59) | 12.92 (10.96-14.72) |

^†^ Genetic risk was defined by tertiles of PRS: low (the lowest tertiles), intermediate (the mid tertiles) and high (the highest tertiles);

^‡^ The HRs were estimated using Cox proportional hazards regression with adjustment for age, sex, BMI, highest education level, family history of cancer, personal medical history (previous cancer diagnoses and chronic obstructive pulmonary disease), the forced expiratory volume in 1 second, and the top ten principal components of ancestry;

^§^ The 5-year absolute risk reduction and 95% CI were generated by drawing 1000 bootstrap samples from the estimation dataset.

**Figure Legends**

Figure E1. Study design and workflow

Figure E2. Flowchart for the calculation of polygenic risk scores (PRSs).

(A) Chinese-specific PRS;

(B) White-specific PRS.

The process of variants filtering is shown on the left, and the final SNP lists as well as their corresponding effects are shown on the right. The largest available GWAS datasets of lung cancer in populations of Chinese descent (13,327 cases and 13,328 controls) and of white descent (29,266 cases and 56,450 controls) were used to re-evaluate the associations and corresponding effects of all the previously reported single-nucleotide polymorphisms associated with lung cancer risk.

Figure E3. The relationship of ethnic-specific polygenic risk scores (PRSs) with incident lung cancer in CKB and UKB cohorts. (Left) Linear relationship between PRS and lung cancer risk was assessed using a restricted cubic spline analysis (A and C); (Right) Participants were divided into ten equal parts (deciles) according to PRS, and the hazard ratio (HR) of each part compared with those at the lowest deciles was shown in the right (B and D). The HRs were estimated using Cox proportional hazards regression with adjustment for age, sex, smoking status, BMI, highest education level, family history of cancer, personal medical history (previous cancer diagnoses and chronic obstructive pulmonary disease), the forced expiratory volume in 1 second, and the top ten principal components of ancestry.

Figure E4. Mediation analysis of smoking on associations between PRS and incident lung cancer risk in the CKB (A) and UKB cohorts (B). The mediation proportion by the mediator was calculated by comparing estimates from models with and without the hypothesized mediator.

Figure E5. Flowchart for the calculation of trans-ancestry polygenic risk scores (PRSs). The process of variants filtering is shown on the left, and the final SNP lists as well as their corresponding effects are shown on the right. A meta-analysis dataset of lung cancer with balanced sample sizes of White (13,793 cases and 14,027 controls from the INTEGRAL-ILCCO OncoArray Project) and Chinese descent (13,327 cases and 13,328 controls) was used to re-evaluate selective candidate variants to construct a trans-ancestry PRS for sensitivity analysis.

Figure E6. Risk of incident lung cancer according to trans-ancestry polygenic risk scores (PRSs) and pack-years of smoking categories in the CKB (A) and UKB cohorts (B). The hazard ratios were estimated using Cox proportional-hazard models with adjustment for age, sex, BMI, highest education level, family history of cancer, personal medical history (previous cancer diagnoses and chronic obstructive pulmonary disease), the forced expiratory volume in 1 second, and the top ten principal components of ancestry.

Figure E7. Absolute risk and risk reduction of incident lung cancer according to pack-years of smoking within each genetic risk category defined by trans-ancestry PRS in the CKB (A) and the UKB (B). Genetic risk was categorized into low (the bottom quintile), intermediate (quintiles 2-4) and high (the top quintile) according to distributions of PRSs. The 5-year absolute risks were standardized for age according to the mean in CKB and UKB synchronously. The HRs were estimated using Cox proportional hazards regression with adjustment for age, sex, BMI, highest education level, family history of cancer, personal medical history (previous cancer diagnoses and chronic obstructive pulmonary disease), the forced expiratory volume in 1 second, and the top ten principal components of ancestry. The 5-year absolute risk reduction and 95% CI were generated by drawing 1000 bootstrap samples from the estimation dataset.

Figure E8. The distributions of trans-ancestry polygenic risk scores (PRSs) in CKB and UKB cohorts. Significantly higher distributions of trans-ancestry PRSs were observed in participants of UKB for whole cohort (P<2.2×10^-16^), nonsmokers (P<2.2×10^-16^), and smokers (P<2.2×10^-16^).

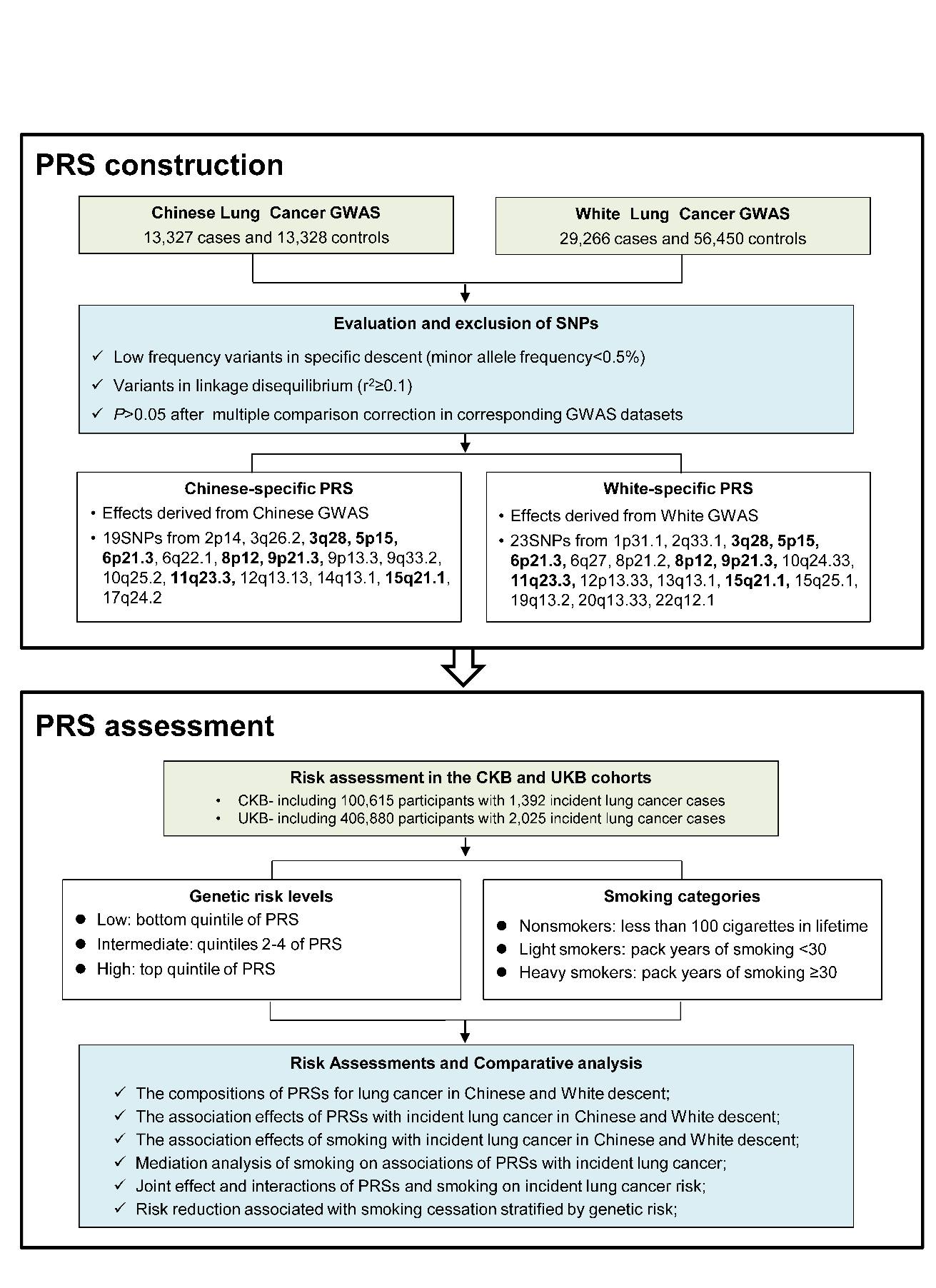

Figure E1

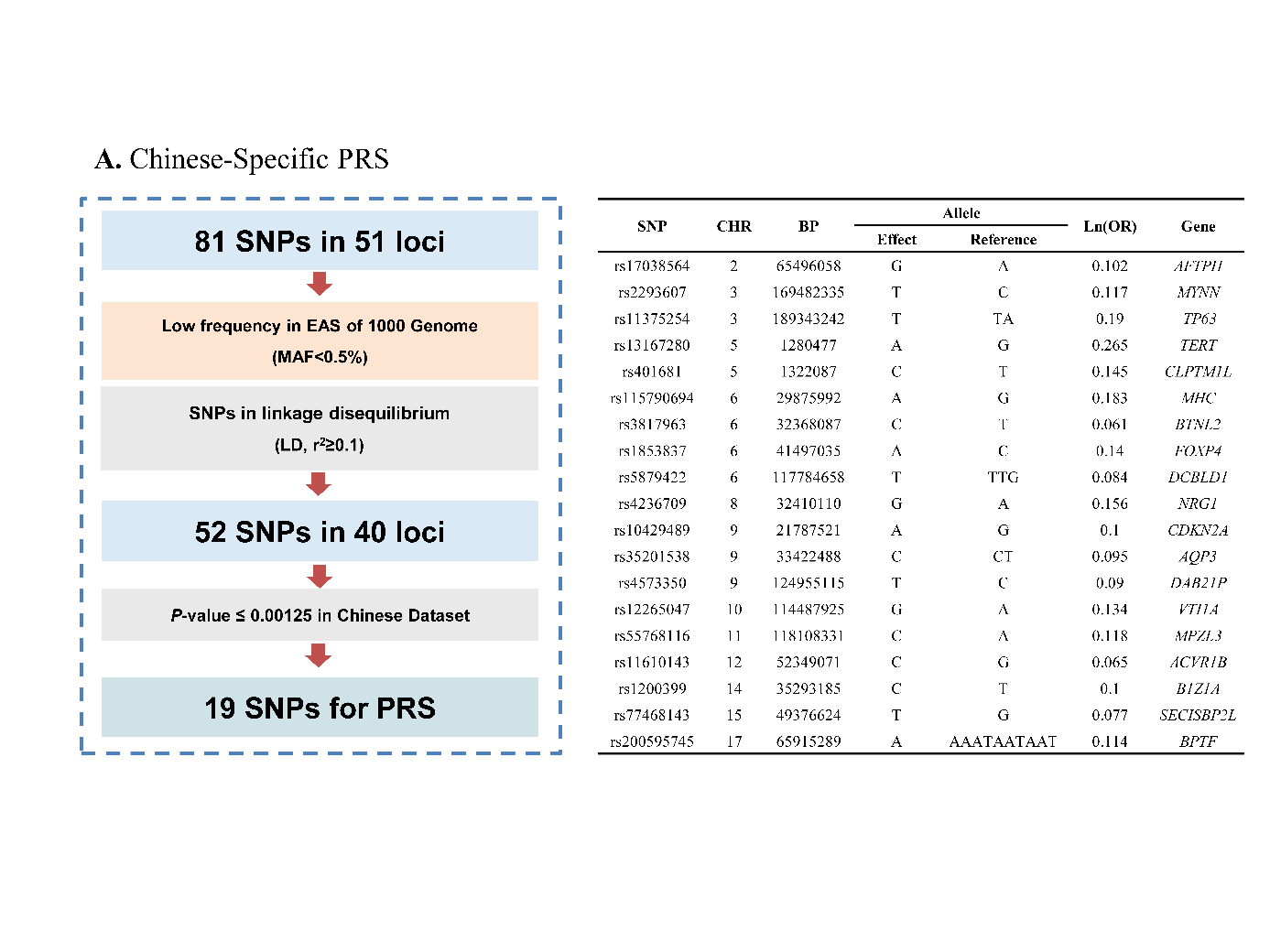

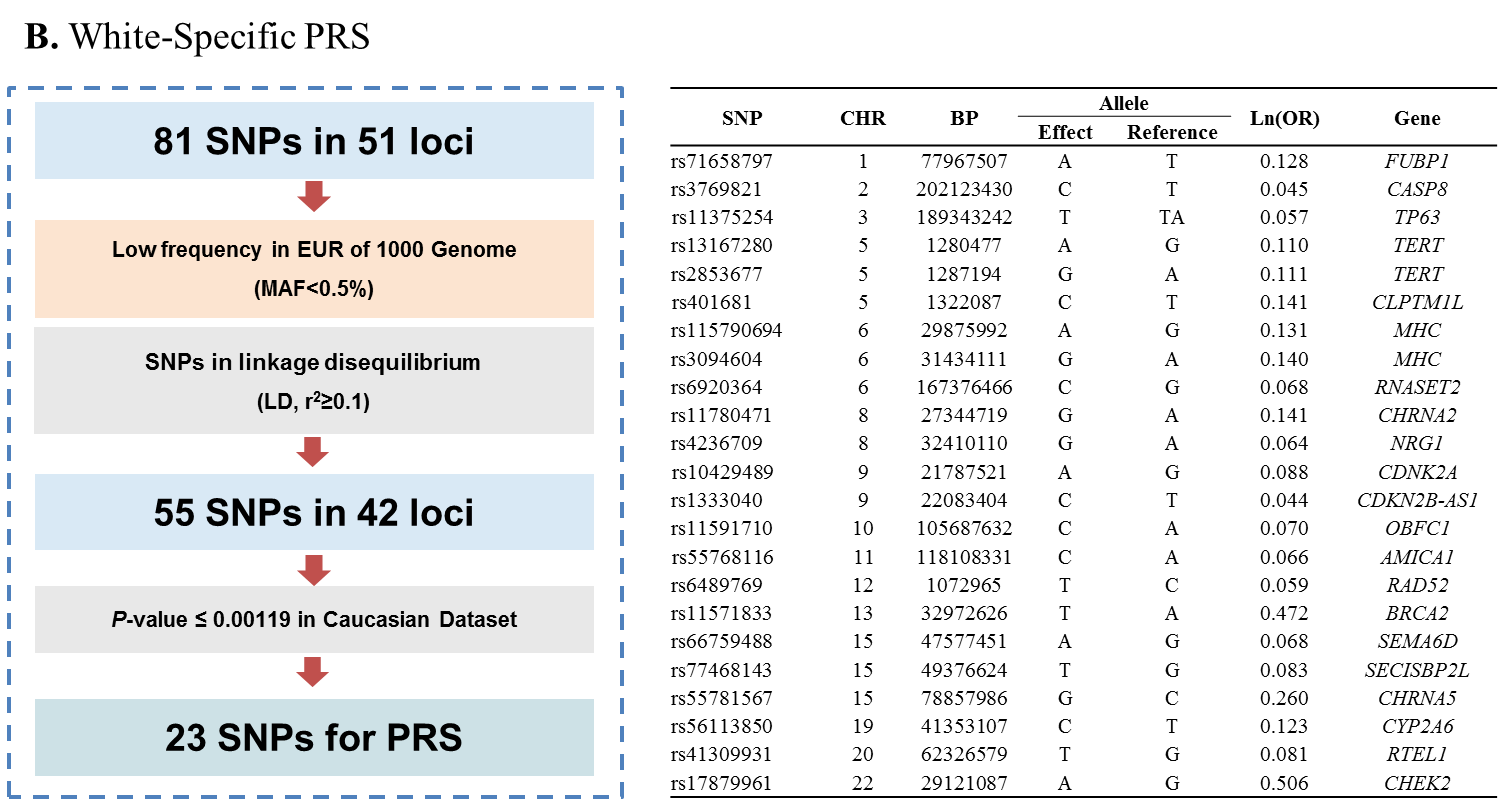

Figure E2

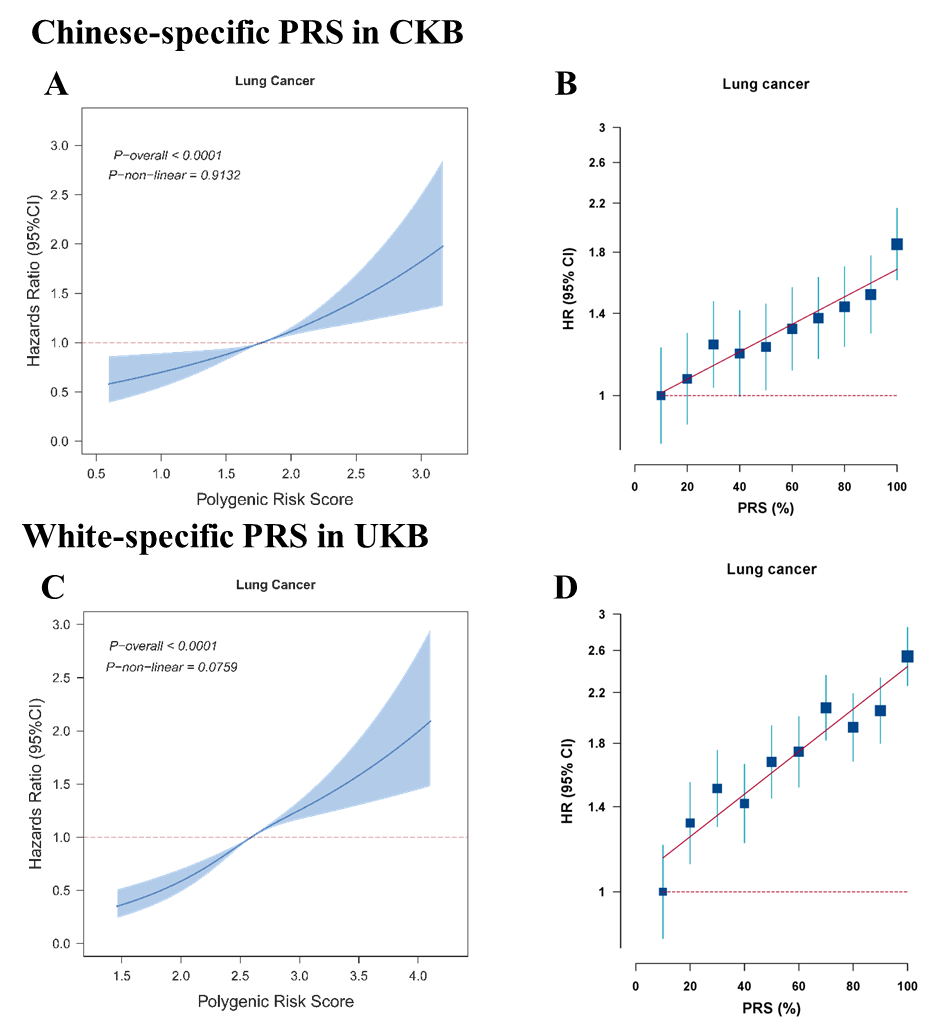

Figure E3

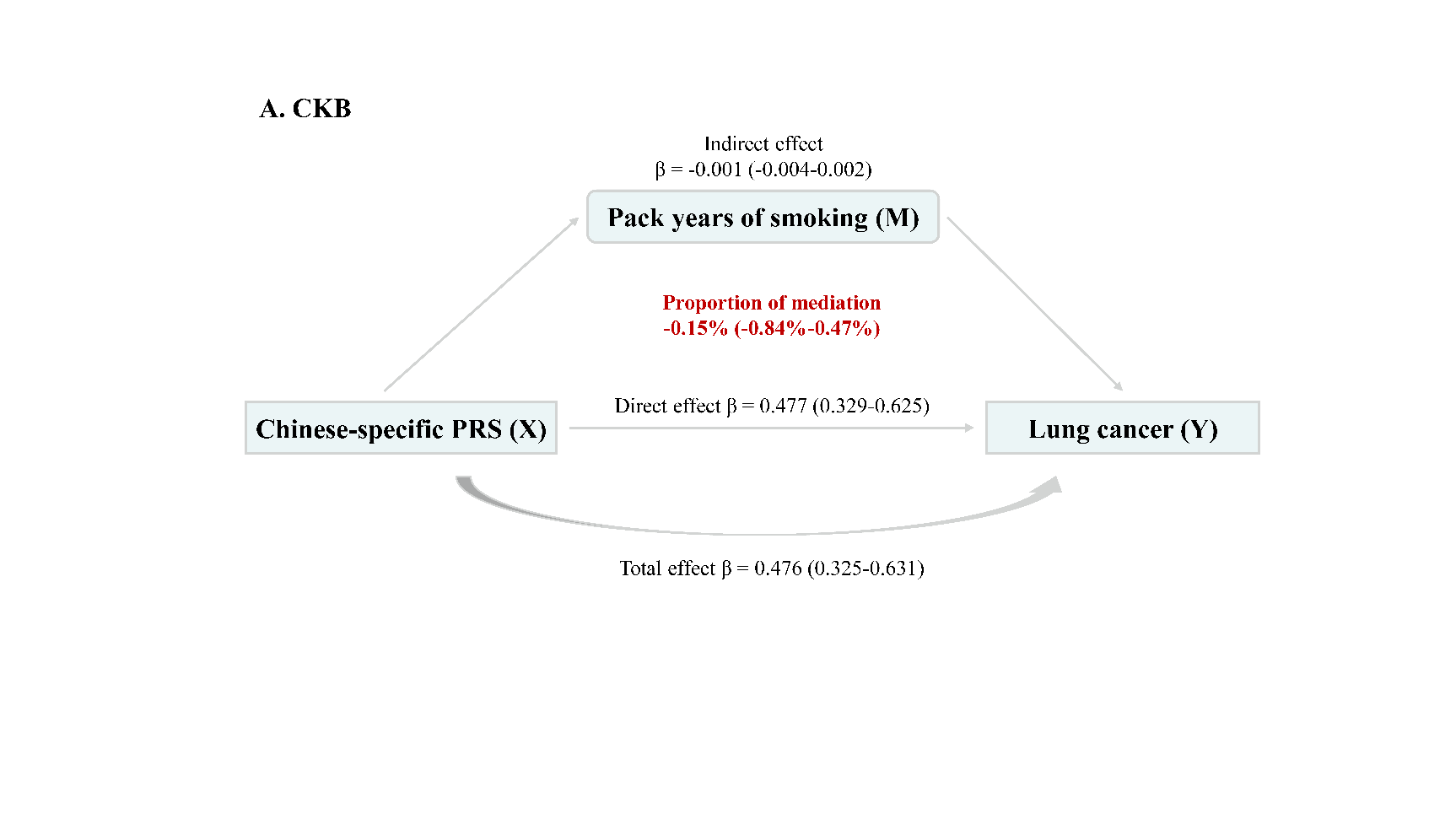

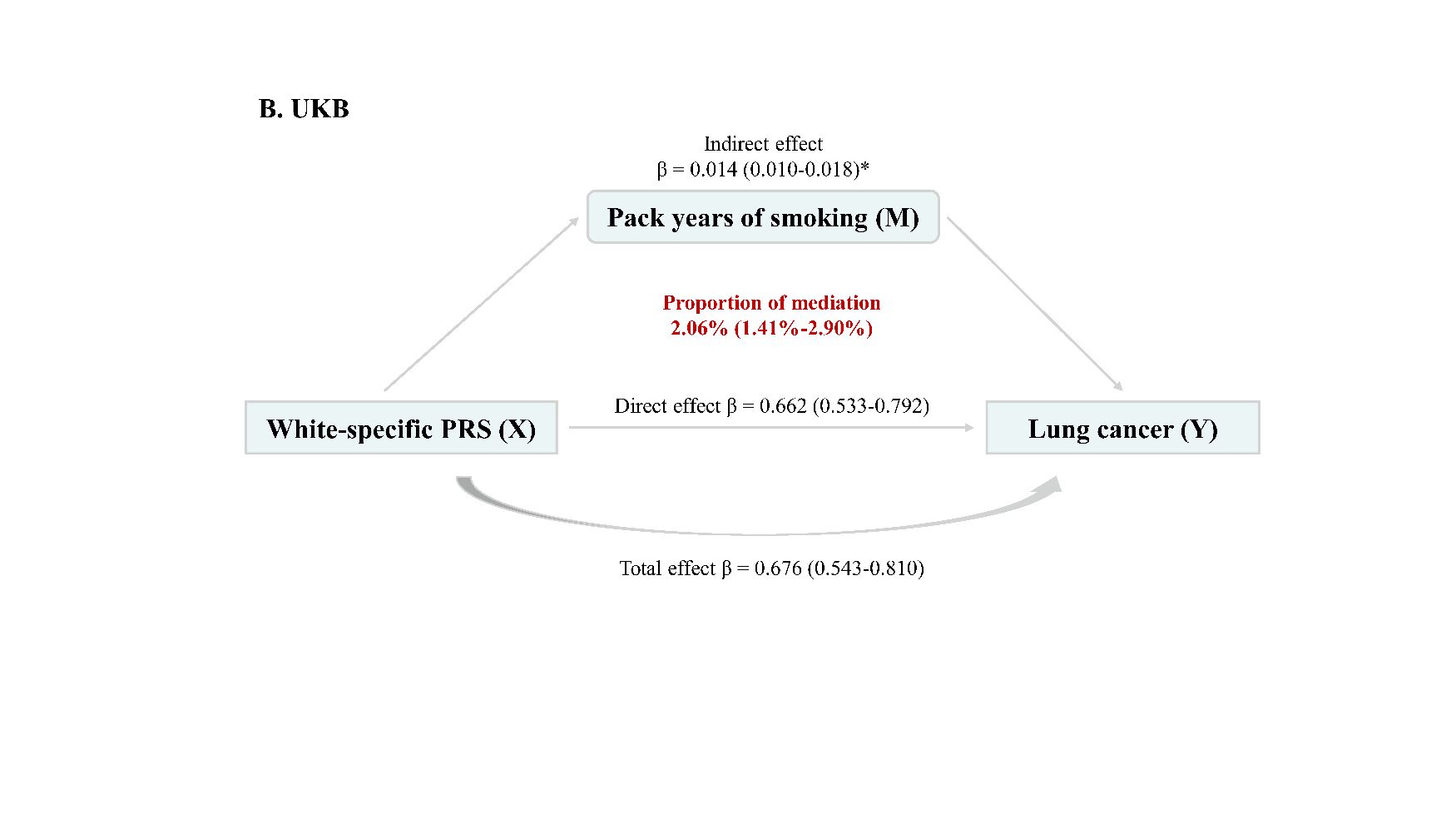

Figure E4

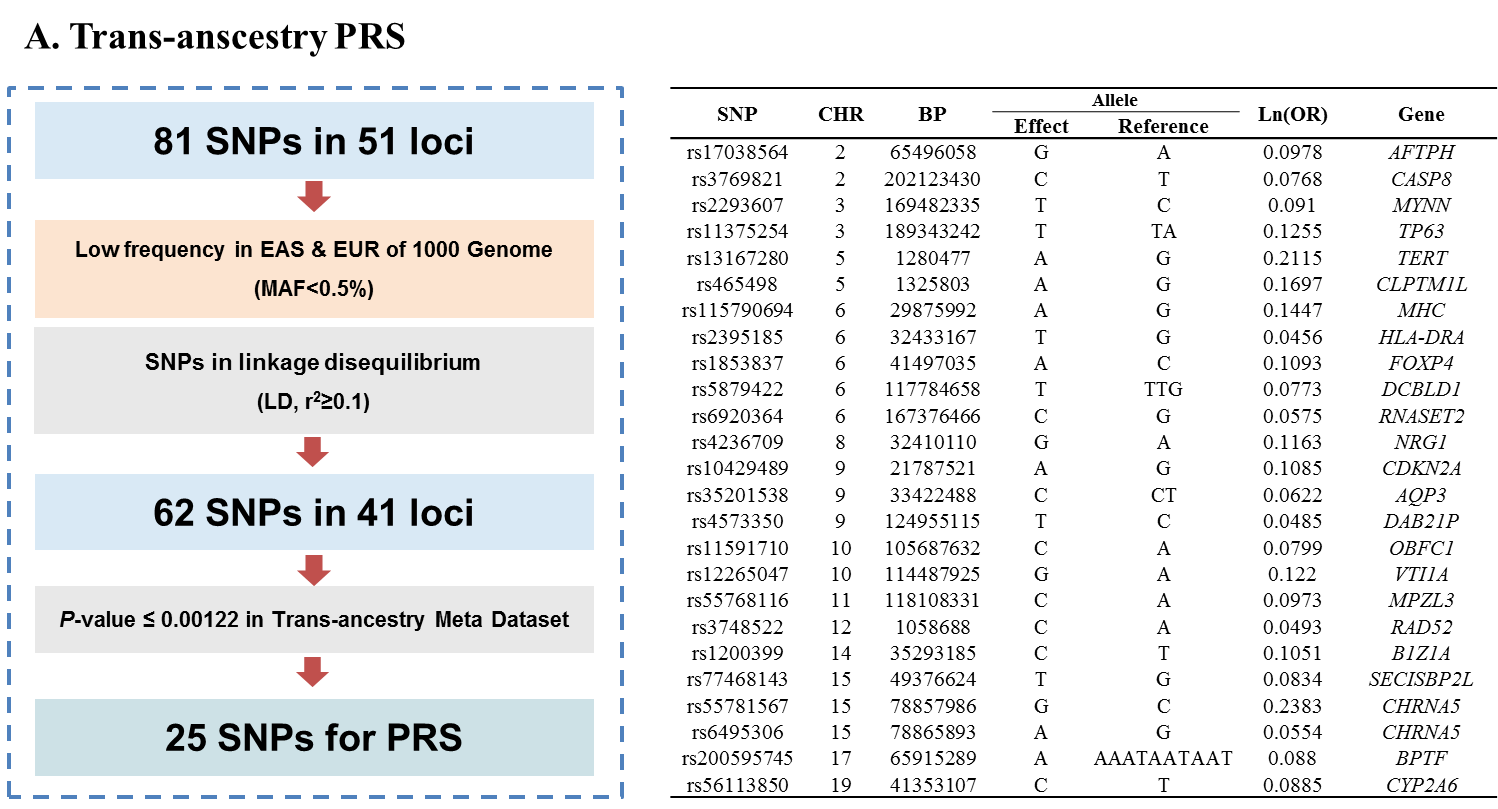

Figure E5

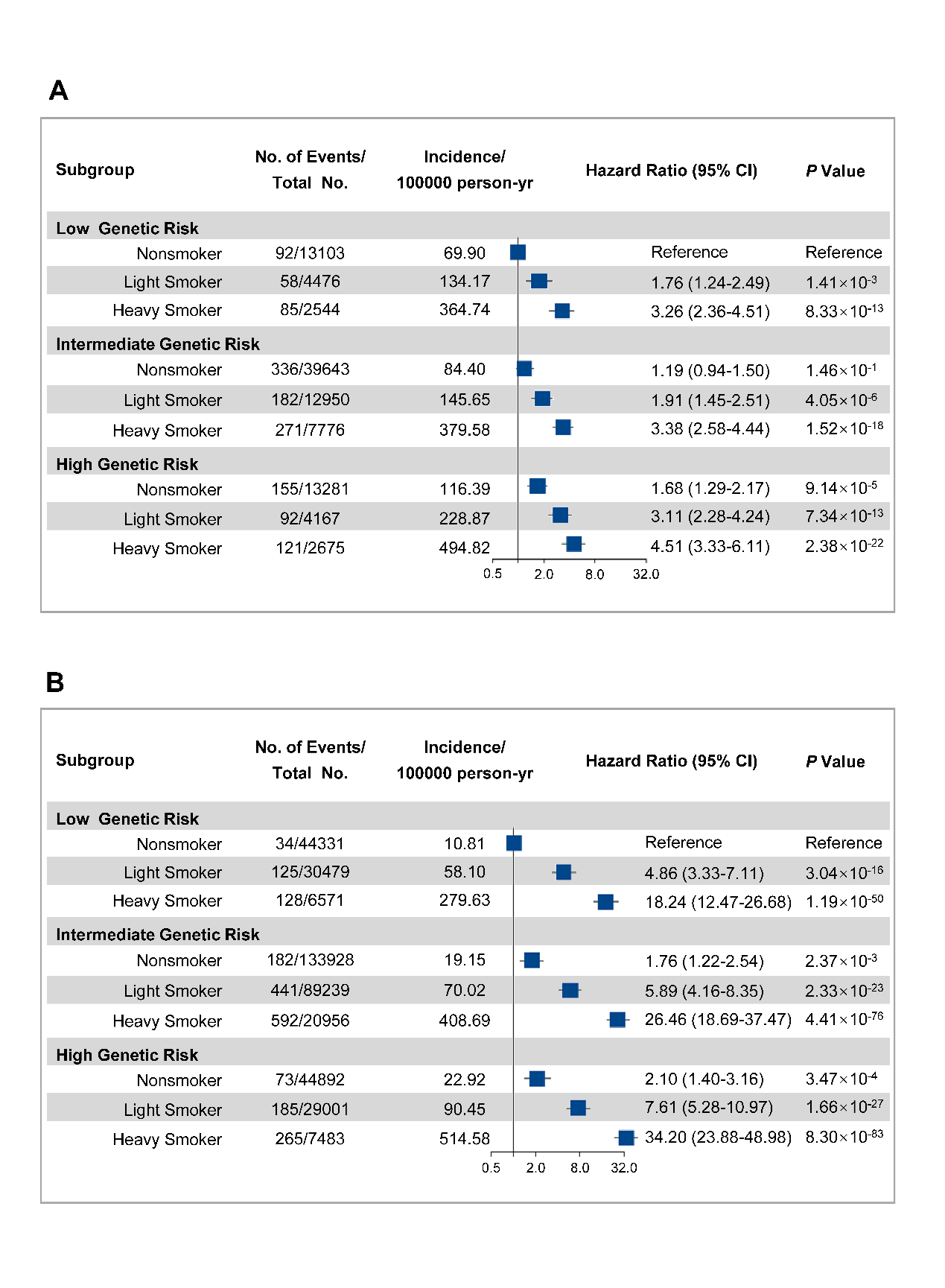

Figure E6

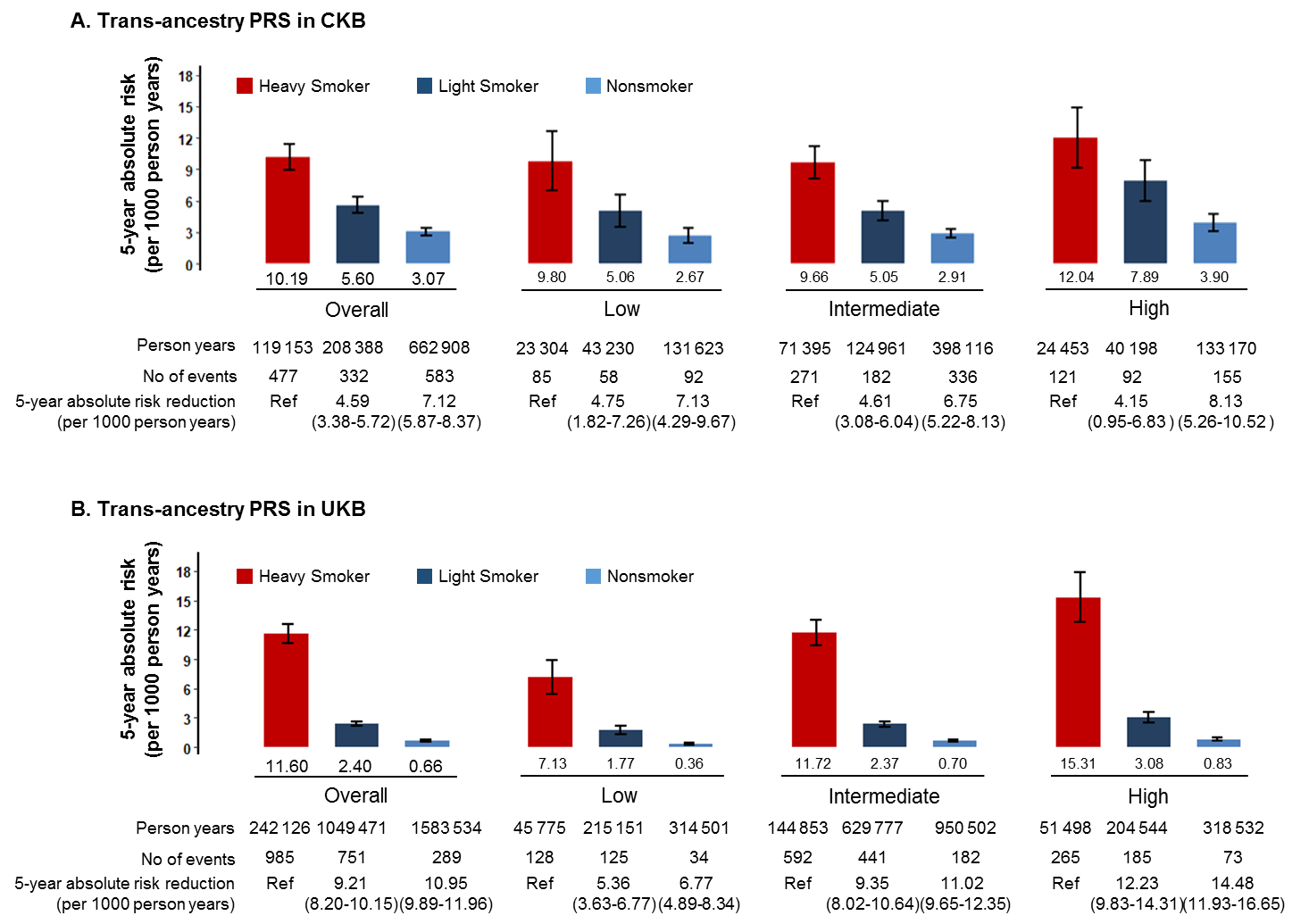

Figure E7

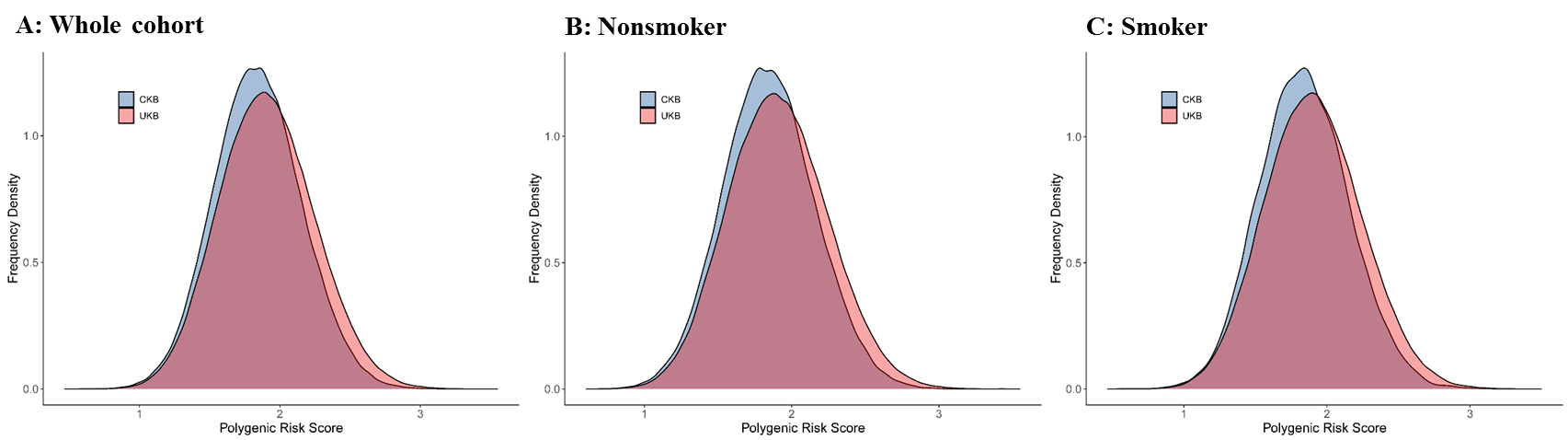

Figure E8
